# MIEHS: A Quantitative Approach to Hub Property Analysis in Brain Networks

**DOI:** 10.1101/2025.05.19.25327906

**Authors:** Hongzhou Wu, Jinming Xiao, Elijah Agoalikum, Benjamin Becker, Stefania Ferraro, Bharat B. Biswal, Benjamin Klugah-Brown, Michael Maes

## Abstract

The human brain relies on dynamic interactions among modular networks, where connector and provincial hubs critically enable information integration. However, existing hub characterization remains predominantly qualitative, overlooking the quantitative contributions of potential hub nodes. To address this gap, we introduce the Multi-Indicator Entropy Hub Score (MIEHS), a quantitative framework integrating six graph-theoretical metrics including betweenness/degree centrality, participation coefficient, within-module betweenness/degree centrality, and clustering coefficient, to holistically evaluate hub properties. We validated MIEHS using benchmark networks, random simulated networks, and resting-state fMRI data from the Midnight Scan Club dataset, demonstrating its accuracy and robustness in hub identification. Our findings reveal that high-score connector hubs predominantly localize within the attention network, while high-score provincial hubs are concentrated in the default mode network (DMN). Gradient mapping further indicates that connector hubs bridge unimodal and transmodal regions, facilitating the transition of information from primary sensory areas to higher-order cognitive regions, whereas provincial hubs primarily support intra-network communication. Additionally, random null model analysis highlights the stability of hub nodes within the DMN and limbic networks. Moreover, to explore clinical implications, we applied Partial Least Squares analysis to the UCLA dataset (HC=110, ADHD=37, BD=40, SCHZ=37), examining the relationship between hub properties and psychiatric symptoms. The results reveal significant associations between hub changes in DMN, SMN, limbic, DAN, and control networks with cognition and behavior. Notably, modifications in hub connectivity impact cognitive flexibility, abstract reasoning, and verbal expression. By bridging quantitative hub analysis with clinical phenotypes, MIEHS provides novel insights of brain network organization and demonstrated a robust tool for elucidating brain functional reconfiguration and functional organization.

## Introduction

The human brain functions through a complex interplay of discrete modular organization and dynamic intermodular interactions, enabling efficient information processing and cognitive flexibility ^1–4^. As large-scale brain network studies continue to evolve ^5^, the role of hub nodes, which are critical regions facilitating communication within and between brain modules ^6,7^, has gained increasing attention. These hubs are essential for maintaining efficient information flow, and their disruption due to neurological disorders or brain injuries can significantly alter cognitive and behavioral functions ^8–10^. Compared to the more diffuse and complex patterns of functional connectivity alterations ^11,12^, focusing on changes in hub properties provides a more direct and interpretable approach to understanding brain mechanisms.

Hub nodes can be broadly categorized into connector hubs, which mediate inter-module communication by linking multiple functional domains^13^, and provincial hubs, which primarily support intra-module communication and local processing ^14^. Traditional approaches to hub identification have relied on either single graph-theoretic measures or a combination of a few metrics, often leading to inconsistencies across studies. While numerous studies have identified the default mode network (DMN) as a central hub in whole-brain networks^15–17^, discrepancies arise when using different evaluation metrics such as degree centrality ^18^, betweenness centrality ^19^, nodal efficiency ^20^, and connection strength ^21^. These inconsistencies are particularly evident in regions such as the thalamus, medial superior frontal gyrus, lateral prefrontal cortex, and supplementary motor area, where different metrics produce varying results regarding their hub status.

The hierarchical nature of brain organization further complicates hub identification, as single-metric approaches struggle to effectively distinguish between inter- and intra-module hubs. While some studies have attempted to refine hub identification by integrating multiple metrics ^17,22,23^, these methods remain largely qualitative, lacking a unified framework for quantifying hub properties and failing to account for the full influence of potential hub nodes within the network ^2^.

To address these challenges, we previously introduced the Multi-Criteria Quantitative Graph Analysis (MQGA) algorithm, which integrates multiple graph-theoretic metrics to systematically evaluate connector and provincial hubs ^24^. This approach showed promise in refining hub identification, but its performance declined as network sparsity decreased and global connectivity increased, particularly in evaluating provincial hubs. Additionally, MQGA lacked validation regarding the theoretical rationale behind its selected metrics and did not fully explore the implications of hub alterations in disease progression. A more robust and adaptable method is required to accurately capture the complexity of hub properties across diverse brain networks.

In this current study, we propose the Multi-Indicator Entropy Hub Score (MIEHS), a novel and comprehensive framework for quantifying hub properties. MIEHS refines provincial hub identification by incorporating within-module betweenness centrality, within-module degree centrality, and clustering coefficient, mitigating the confounding influence of global connectivity on local network structures. Additionally, we introduce an entropy-based weighting system, which assigns different informational values to each graph-theoretic metric, ensuring a more balanced and accurate evaluation of hub nodes. This methodological advancement allows for a more precise differentiation between connector hubs and provincial hubs, improving the reliability of hub identification across varying network configurations.

## Materials and Methods

### Multi-Index Entropy Hub Score (MIEHS)

To quantitatively evaluate hub properties, we developed the MIEHS, integrating multiple graph-theoretic indicators to enhance the accuracy of hub identification. The connector hub (*CON*) score is computed using three fundamental graph theory metrics: betweenness centrality (BC), degree centrality (DC), and participation coefficient (PC). DC directly measures a node’s overall connectivity, allowing for the initial identification of highly influential candidates. BC captures “bridge” nodes that facilitate information integration across the network, while PC highlights cross-modular connectivity, a defining characteristic of connector hubs.

Since increased inter-modular connectivity, or reduced network sparsity, can diminish the predictive accuracy of provincial hub (*PRO*) identification, we refine the *PRO* score using within-module betweenness centrality (wBC), within-module degree centrality (wDC), and clustering coefficient (CC). Normalizing wDC and wBC as Z-scores mitigates the confounding effects of inter-modular connections. Additionally, CC quantifies the clustering tendency of provincial hubs within their local network, further enhancing the accuracy of provincial hub identification.

To more precisely capture the graph properties of each node, MIEHS computations are based on weighted network measures rather than binary networks, ensuring a more nuanced representation of connectivity patterns ^25^. Detailed calculation methods for these indicators are provided in the **Supplementary Methods**.

Before performing calculations, we applied a transformation to betweenness BC, DC, wBC, and wDC by computing their square roots. This adjustment enhances the discrimination between nodes and improves the normality of the resulting distributions. Since wBC and wDC contain negative values, we took the square root of their absolute values and reassigned the corresponding negative signs to maintain their directional properties.

Following this transformation, we constructed two feature vectors: *X_con_* for computing the *CON* score and *X_pro_* for computing the *PRO* score. The feature vectors are defined as follows:

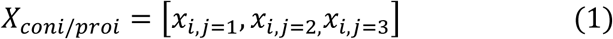

Where *x*_*i*,1_ = *BC*_*i*_, *x*_*i*,2_ = *DC*_*i*_, and *x*_*i*,3_ = *PRO*_*i*_ in the *X_con_*, and *x*_*i*,1_ = *wBC*_*i*_, *x*_*i*,2_ = *wDC*_*i*_, and *x*_*i*,3_ = *CC_i_* in the *X_proi_*. For each subject/network, *X* is a matrix with *i* representing different ROIs and *j* representing three different graph indicates. To enhance the sensitivity of the MIEHS algorithm in identifying hub nodes, we accounted for the varying information weights provided by different graph theory metrics. Entropy weights were calculated separately for *X_con_* and *X_pro_* respectively:

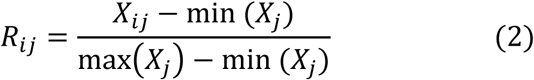

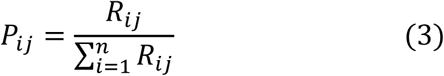

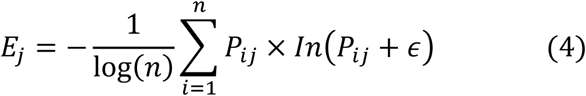

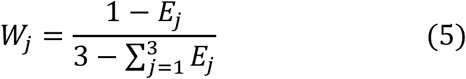

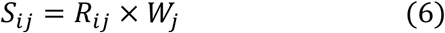

Formula 2 to Formula 6 show the process of entropy weight processing. *R*_*ij*_ is the standardized *X_con/pro_* matrix, *P_ij_* is the corresponding weight matrix, *E_j_* is the information entropy corresponding to each indicator, *W*_*j*_ is the weight of each indicator, and *S_ij_* represents the comprehensive score matrix after the standardized graph theory indicator is assigned with the corresponding weight. In order to further eliminate the impact of different dimensions and ensure that the result conforms to the normal distribution, we normalized the comprehensive score *S*_*ij*_:

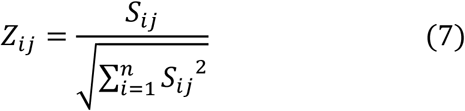

We then calculated the Euclidean distances *D*^+^_*i*_ and *D*^−^_*i*_ between the attribute vector and Z^+^ and Z^−^ of each ROI of different subjects:

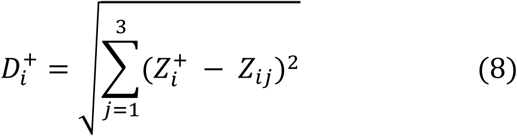

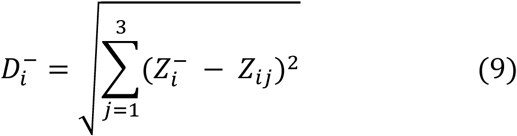

Where Z^+^ = [max{Z_*i*1_}, max{Z_*i*2_}, max{Z_*i*3_}] and Z^−^ = [min{Z_*i*1_}, min{Z_*i*2_}, min{Z_*i*3_}]. Finally, the *con* and *pro* indicators of each ROI by using *D*^+^ and *D*^−^ were calculated:

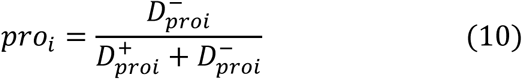

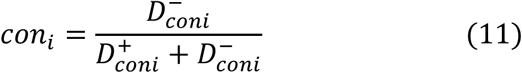

*D_coni_* represent the result calculated by using BC, DC and PC indicators through formulas 1 to 9, while *D_proi_* represent the result calculated by using wBC, wDC and CC through formulas 1 to 9.

### Simulation network analysis

To preliminarily validate the effectiveness of the MIEHS algorithm, we applied it to two benchmark networks and 100 randomly generated complex networks. For the benchmark networks, we first constructed a 12-node network with two sub-networks, where one node was designated as a connector hub and two as provincial hubs. We applied MIEHS to this network and ranked the nodes based on their *CON* and *PRO* scores. Next, we tested MIEHS on *Zachary’s karate club* network (34 nodes), a widely used benchmark in network analysis^26^, and explored the relationship between hub scores and diffusion time using a Simplified Susceptible-Infected (SSI) model ^27^. The specific SI score of ‘*Zachary’s karate club*’ network and detailed information on the simplified SI model were provided in the **Supplementary Table S1** and **Supplementary methods**.

To further assess MIEHS in random networks and evaluate its performance under varying sparsity conditions, we generated 100 randomly connected networks, each consisting of 100 nodes and 5 modules with designated connector hubs, provincial hubs, and normal nodes. We examined the distribution of *CON* and *PRO* scores across different sparsity thresholds and computed Pearson correlations between hub scores and node labels. Additionally, we conducted a vulnerability analysis, measuring the reduction in network efficiency upon node deletion. Notably, for PRO vulnerability analysis, we considered only the efficiency reduction within the affected sub-network, performing correlation calculations between *PRO* scores and the local efficiency loss. The construction details of the synthetic networks are described in ^24^.

### fMRI data preprocessing and functional connectivity matrix construction

To further validate and apply MIEHS, we utilized resting-state fMRI data from the Midnight Scan Club (MSC) dataset and the University of California, Los Angeles (UCLA) Consortium for Neuropsychiatric Phenomics (CNP) dataset ^28–30^. The MSC dataset, which provides extensive long-term resting-state fMRI scans for individual subjects, is well-suited for validating MIEHS’s stability. The UCLA dataset, which includes data from healthy controls (HC), bipolar disorder (BD), attention deficit hyperactivity disorder (ADHD), and schizophrenia (SCHZ) patients, allows us to evaluate MIEHS’s performance across multiple clinical conditions.

The MSC dataset consists of 10 subjects (5 females, aged 24–34), each undergoing ten 30-minute resting-state fMRI scans. Based on prior findings, one subject (MSC08) was excluded due to abnormal outcomes^28^ and **Supplementary Figure S1**.

The UCLA dataset includes 272 participants, comprising 130 healthy controls (HC), 49 BD, 43 ADHD, 39 schizophrenia (SZ), and 11 schizoaffective disorder (SZAD) patients. Since SZ and SZAD groups exhibit overlapping characteristics, they were merged into a single SCHZ category. Following Kebets et al. (2019) exclusion criteria, we retained 110 HC, 37 ADHD, 40 BD, and 37 SCHZ patients ^31^. Further demographic and clinical details are provided in **Supplementary Tables S2** and **S3**. Preprocessed BOLD time series were standardized to the 32k fs_LR surface and parcellated into 400 regions based on the Yeo7 400-ROI atlas ^32,33^. Functional connectivity matrices (400×400) were constructed using Pearson’s correlation coefficients after smoothing the time series with 2 mm Gaussian kernels. Full preprocessing details are available in the **Supplementary Methods**.

### Intra-class correlation coefficient (ICC) analysis

To assess the stability of CON and PRO scores, we performed intra-class correlation coefficient (ICC) analysis on the remaining nine MSC subjects across their ten resting-state scans. ICC values were calculated using a one-way random effects model, measuring absolute agreement across scans. We then computed Pearson correlations between average CON and PRO scores and ICC values across all 400 ROIs, providing insight into the reliability of MIEHS in repeated measures.

### Gradient Mapping

We mapped hub scores onto functional gradients to further validate the computational accuracy of the MIEHS approach and examine its relationship with gradient distributions. Using the Yeo7 template, we segmented the functional gradient distribution derived in studies by *Margulies et al.* and *Markello et al.* ^34,35^, mapping the CON (and PRO) values onto the Gradient 1 and Gradient 2.

### Random null model analysis

To test the statistical significance of hub scores, we employed a random null model that preserves node degree while randomizing connectivity patterns. Based on our simulation network analyses and ICC findings, networks with stable sparsity threshold 0.25 were selected for randomization. We generated 1,000 random networks per original network by randomly swapping edges 100 times the total number of connections to achieve sufficient randomness.

To explore the distribution patterns of CON and PRO within brain networks, we constructed two levels of random networks: (1) *Global Exchange Model*: Randomly swaps edges across the entire network, highlighting the importance of nodes within the global topology. (2) *Modular Feature Preservation Exchange*: Swaps edges within or between specific modules, emphasizing functional roles within localized subnetworks. This approach allowed us to assess hub stability and significance under different structural constraints.

### Partial Least Squares (PLS) analysis

To examine the relationship between hub scores and behavioral phenotypes, we performed Partial Least Squares (PLS) analysis on 52 behavioral measures and hub scores from 224 participants, following the approach of ^31^. PLS is a multivariate technique that extracts latent components (LCs), which represent optimal linear combinations of hub scores and behavioral data ^36,37^. Each LC revealed distinct patterns of hub-behavior associations.

To quantify individual contributions, we projected each participant’s hub scores and behavioral data onto salience maps, computing composite hub and behavioral scores for each LC. The number of significant LCs was determined using 1,000 permutation tests, applying False Discovery Rate (FDR) correction (q < 0.05) to top five LCs.

To further dissect these LCs, we calculated Pearson correlations between: (1) Behavioral measures and LC-derived behavioral composite scores. (2) Hub scores and LC-derived hub composite scores. High positive or negative correlations indicated key behavioral and hub contributors within each LC. To ensure statistical robustness, we employed bootstrapping with 1,000 resampled datasets, computing Z-scores for correlation coefficients, which were converted to p-values with additional FDR correction (q < 0.05). This analysis provided a comprehensive mapping of hub-behavior relationships, offering insight into how altered hub properties influence cognition, executive function, and psychiatric traits.

## Results

### Benchmark and random network verification

The overall MIEHS process is illustrated in Figure 1A, where six graph-theoretic indicators are combined across different dimensions to compute *CON* and *PRO* scores. Entropy weights were applied to balance the contribution of these indicators, enabling a more precise and nuanced hub score computation.

**Figure 1.**
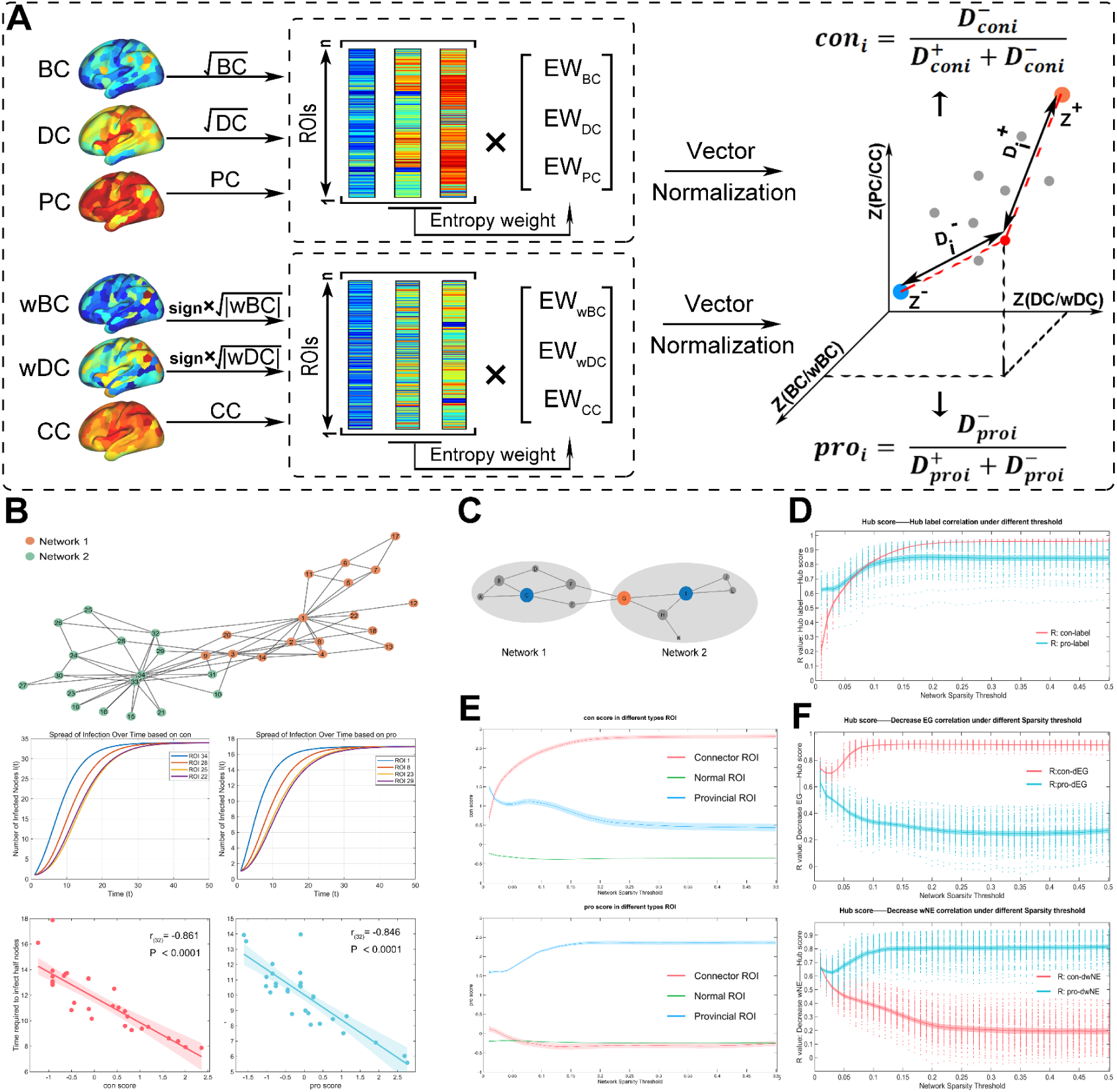
MIEHS and Simulation Verification. (A) Visualization results of MIEHS calculation process. (B) ‘*Zchary’s karate club*’ benchmark network and SSI model verification. The middle part shows the time-diffusion node results of four typical ROIs, and the bottom part shows the *Pearson* correlation results of the time t taken to diffuse half of the nodes and the hub score. Note: In the propagation simulation of PRO, only propagation to all nodes in the sub-network is considered. (C) 12-node simple benchmark network. (D) Hub score and node label correlation results. (E) Distribution of hub scores of different types of nodes. (F) The correlation between reduced network efficiency after deleting the corresponding node and hub score.

The Benchmark network analysis is shown in Figure 1B. The four highlighted nodes (1st, 11st, 21st, and 31st) were selected based on their ranked *CON* and *PRO* scores, sorted from highest to lowest. This observation was further supported by a significant negative correlation between hub scores and the time needed to reach half of the network nodes, as shown in the lower part of Figure 1B. The complete ranking of hub scores for all nodes in this benchmark network is provided in **Supplementary Table S1**.

To further verify the validity of MIEHS, we applied it to a simplified 12-node network, as depicted in Figure 1C depicts verification of the validity of MIEHS. Both MQGA and MIEHS produced identical *CON* rankings, confirming their robustness in identifying connector hubs. However, the *PRO* rankings in MIEHS more accurately distinguished provincial hub nodes. For example, in the new ranking, node B was assigned fourth place instead of node G, which is a more reasonable placement. Node G had only two intra-module connections within Network 2, whereas node B had three connections. Additionally, nodes B, A, and C formed a triangular structure, resulting in a higher clustering coefficient, further reinforcing node B’s role as a provincial hub. The detailed hub scores for this benchmark network are presented in Table 1.

**Table 1.**
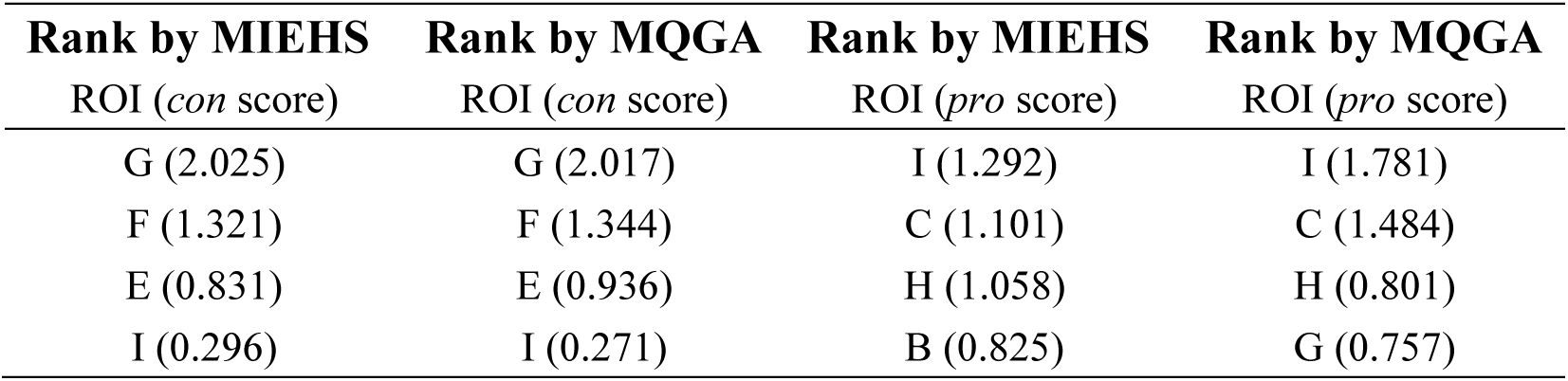
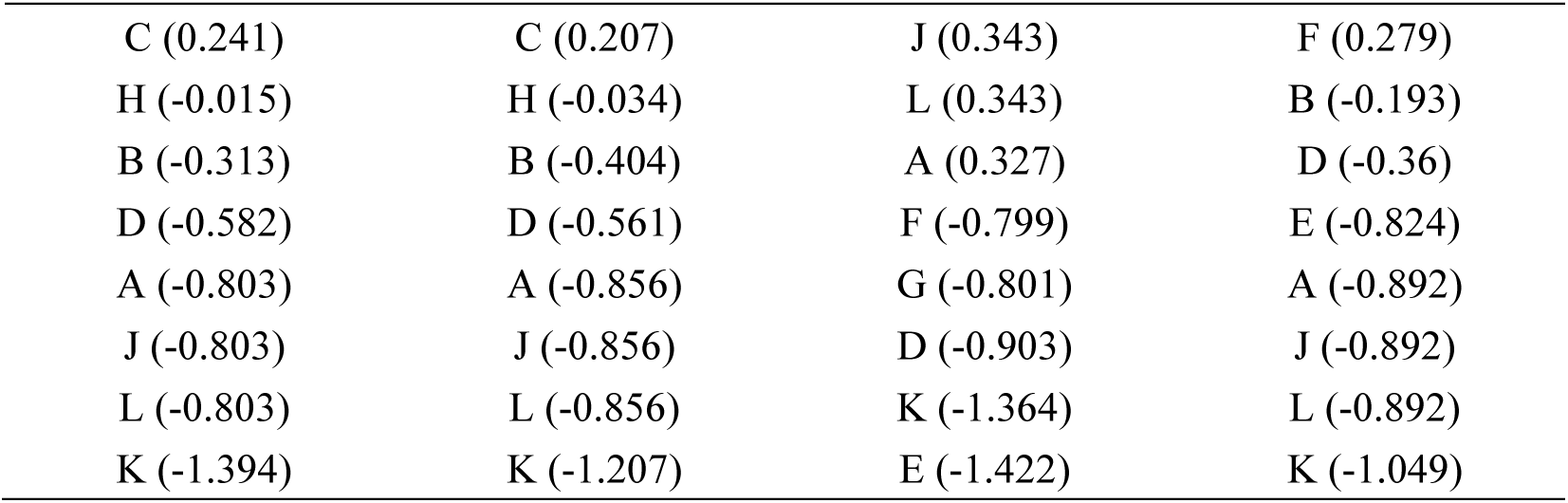
Performance of MQGA and MIEHS on 12-node network

| Rank by MIEHS | Rank by MQGA | Rank by MIEHS | Rank by MQGA |
| --- | --- | --- | --- |
| ROI (*con* score) | ROI (*con* score) | ROI (*pro* score) | ROI (*pro* score) |
| G (2.025) | G (2.017) | I (1.292) | I (1.781) |
| F (1.321) | F (1.344) | C (1.101) | C (1.484) |
| E (0.831) | E (0.936) | H (1.058) | H (0.801) |
| I (0.296) | I (0.271) | B (0.825) | G (0.757) |
| C (0.241) | C (0.207) | J (0.343) | F (0.279) |
| H (-0.015) | H (-0.034) | L (0.343) | B (-0.193) |
| B (-0.313) | B (-0.404) | A (0.327) | D (-0.36) |
| D (-0.582) | D (-0.561) | F (-0.799) | E (-0.824) |
| A (-0.803) | A (-0.856) | G (-0.801) | A (-0.892) |
| J (-0.803) | J (-0.856) | D (-0.903) | J (-0.892) |
| L (-0.803) | L (-0.856) | K (-1.364) | L (-0.892) |
| K (-1.394) | K (-1.207) | E (-1.422) | K (-1.049) |

To assess MIEHS in more complex network structures, we applied it to 100 randomly generated networks, each consisting of 100 nodes and 5 modules. Figures 1D to 1F illustrate the validation results, which include correlation analysis between hub scores and node labels, distribution patterns of CON and PRO scores among different node types, and vulnerability testing at both global and local levels. MIEHS exhibited a significantly higher correlation between hub scores and node labels, improving from R = 0.6 in MQGA to R = 0.8 ∼ 0.9. This correlation remained stable when network sparsity exceeded 0.2, demonstrating the robustness of the MIEHS algorithm. When analyzing the distribution of hub scores among different node types, MIEHS effectively distinguished between connector hubs, provincial hubs, and ordinary nodes. Specifically, connector hubs exhibited the highest CON scores, followed by provincial hubs, while ordinary nodes had the lowest scores. Similarly, provincial hubs showed the highest PRO scores, with connector hubs scoring lower and ordinary nodes ranking the lowest.

In the vulnerability analysis, nodes with higher CON scores had a more significant impact on global network efficiency, whereas nodes with higher PRO scores had a stronger influence on local network efficiency. These results contrast with the MQGA algorithm, where PRO scores failed to accurately capture the role of provincial hub nodes. Although MQGA performed comparably to MIEHS in identifying CON scores during the vulnerability test using MSC data, the correlation between MQGA’s PRO scores and the reduction in local network efficiency diminished as network sparsity increased. Furthermore, the relationship between CON scores and the reduction in local efficiency was nearly identical to that of PRO scores, suggesting that MQGA was unable to distinguish provincial hubs from connector hubs effectively. These findings highlight the superior accuracy and stability of MIEHS in differentiating hub roles and evaluating network vulnerabilities. The complete validation results comparing MQGA with MIEHS, as well as additional analyses using the MSC dataset, are presented in **Supplementary Figure S2**.

### Comprehensive analysis based on MSC dataset

Figure 2 provides further validation of the hub scores derived from the MSC dataset and their relationship with other network metrics. The stability analysis in Figure 2A demonstrates that hub scores remain highly consistent across network sparsity thresholds ranging from 0.01 to 0.5. Notably, *PRO* scores exhibit slightly higher stability than CON scores. This difference arises because the calculation of CON scores incorporates more global variables, leading to lower intra-class correlation coefficient (ICC) values. In contrast, *PRO* scores rely on fewer variables, resulting in a slightly higher ICC. The lower panel of Figure 2A further supports this finding, where a correlation analysis between average hub scores and ICC values reveals a significant positive correlation for both *CON* and *PRO* scores, with CON scores showing a stronger association. This indicates that nodes with higher hub scores exhibit greater stability across scans compared to those with lower scores.

**Figure 2.**
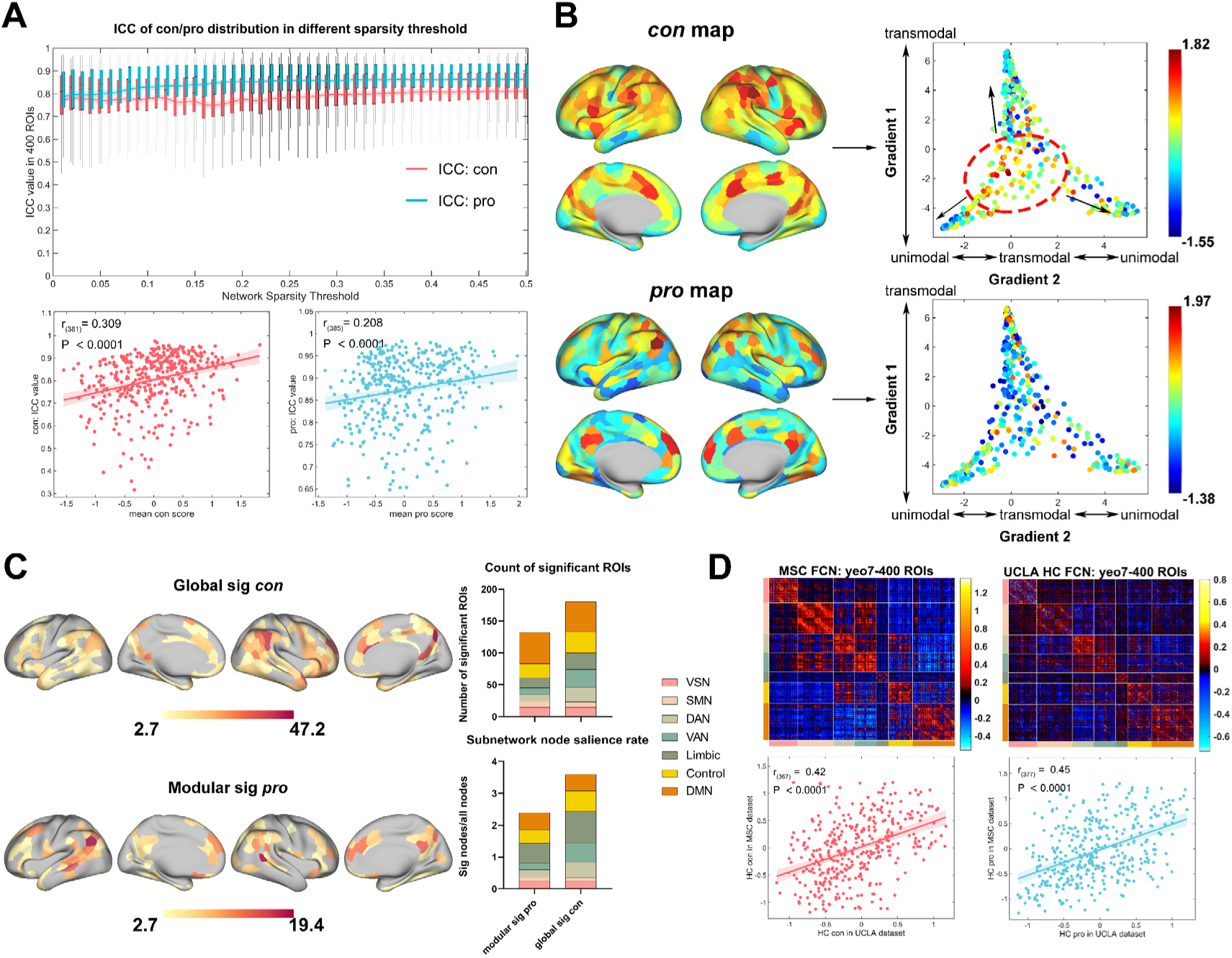
Verification and analysis of MSC dataset. (A) ICC analysis and its correlation results with hub score (sparsity threshold = 0.25). (B) Mean hub scores whole brain mapping and functional gradient mapping. (C) Random Null Model Z-hub scores vs. Hub scores and significant Z-score mapping (FDR correction, q < .01). (D) Upper, average resting-state functional connectivity of healthy subjects in the MSC dataset and UCLA dataset. Lower, correlation of hub scores among healthy subjects across datasets. VSN: Visual Network; SMN: Somatomotor Network; DAN: Dorsal Attention Network; VAN: Ventral Attention Network; Limbic: Limbic Network; Control: Control Network; DMN: Default Mode Network;

The network affiliations of the top 10% of nodes with the highest average hub scores, shown in the left panel of Figure 2B and detailed in **Supplementary Table S4**, reveal distinct spatial distributions for connector and provincial hubs. ROIs with high CON scores predominantly localize within the attention network, whereas ROIs with high PRO scores are primarily found within the ***DMN***. Mapping the average hub scores onto the cortical gradient distribution further illustrates these differences. Nodes with higher CON scores tend to cluster at the transition between *unimodal and transmodal cortical gradients*, suggesting that connector hubs facilitate information flow across hierarchical processing regions. In contrast, PRO scores display a more widespread distribution, with high PRO-scoring ROIs found across both unimodal and transmodal gradients, indicating their role in local information processing.

Figure 2C further supports these observations by showing the validation of *CON* hub scores using a global random null model. The results highlight the central role of connector hubs in the global network, as evidenced by the significant presence of CON-scoring ROIs. The DMN network contains the highest number of significant ROIs, with 29 ROIs demonstrating significance in both *CON* and *PRO* scores, as detailed in **Supplementary Table S5**. However, considering the potential influence of network size on null model outcomes, we normalized the number of significant ROIs by dividing by the total number of ROIs in each respective network. Interestingly, despite having fewer ROIs, the Limbic network exhibited the highest significance rate, with all its ROIs showing significance in the global random null model validation for CON scores. For further evaluation of the generalizability of MIEHS, as shown in Figure 2D, despite different preprocessing process and cross dataset of HC groups between the MSC and UCLA datasets, the average hub scores for both *CON* and *PRO* nodes in the HC groups showed significant positive correlations across datasets, with *R_con_* = 0.42 and *R_pro_* = 0.45. Notably, the cross-dataset correlation for PRO scores was higher than that for CON scores, further supporting the robustness of provincial hub identification.

#### PLS analysis based on UCLA dataset

To investigate the relationship between whole-brain hub scores (*CON* and *PRO*) and behavioral traits, we conducted PLS analysis on 52 behavioral measures from 224 participants in the UCLA dataset across different diagnostic categories. The covariance explained by each LC in the PLS analysis is illustrated in **Supplementary Figure S3**.

For *CON* scores, LC1, LC2, LC4, and LC5 passed permutation tests with FDR correction (q < 0.05), whereas for *PRO* scores, LC1, LC2, and LC4 passed the same tests. Given that LCs beyond LC1 and LC2 explained only limited covariance in control analyses (less than 10%), our primary focus remained on LC1 and LC2, as they accounted for the most substantial variance in the data.

LC1 explained 26% of the covariance for *CON* scores and 25% for *PRO* scores respectively, indicating its strong contribution to hub-related variability. As depicted in Figures 3A and 3F, the distributions of composite scores for hub metrics (*CON* and *PRO*) and behavioral measures were highly similar, suggesting a consistent pattern across both hub classifications. Both hub composite scores were significantly positively correlated with behavioral composite scores (*r_con_* = 0.707, *r_pro_* = 0.696, *p_(con/pro)_* < 0.001), highlighting a strong association between hub organization and behavioral traits.

**Figure 3.**
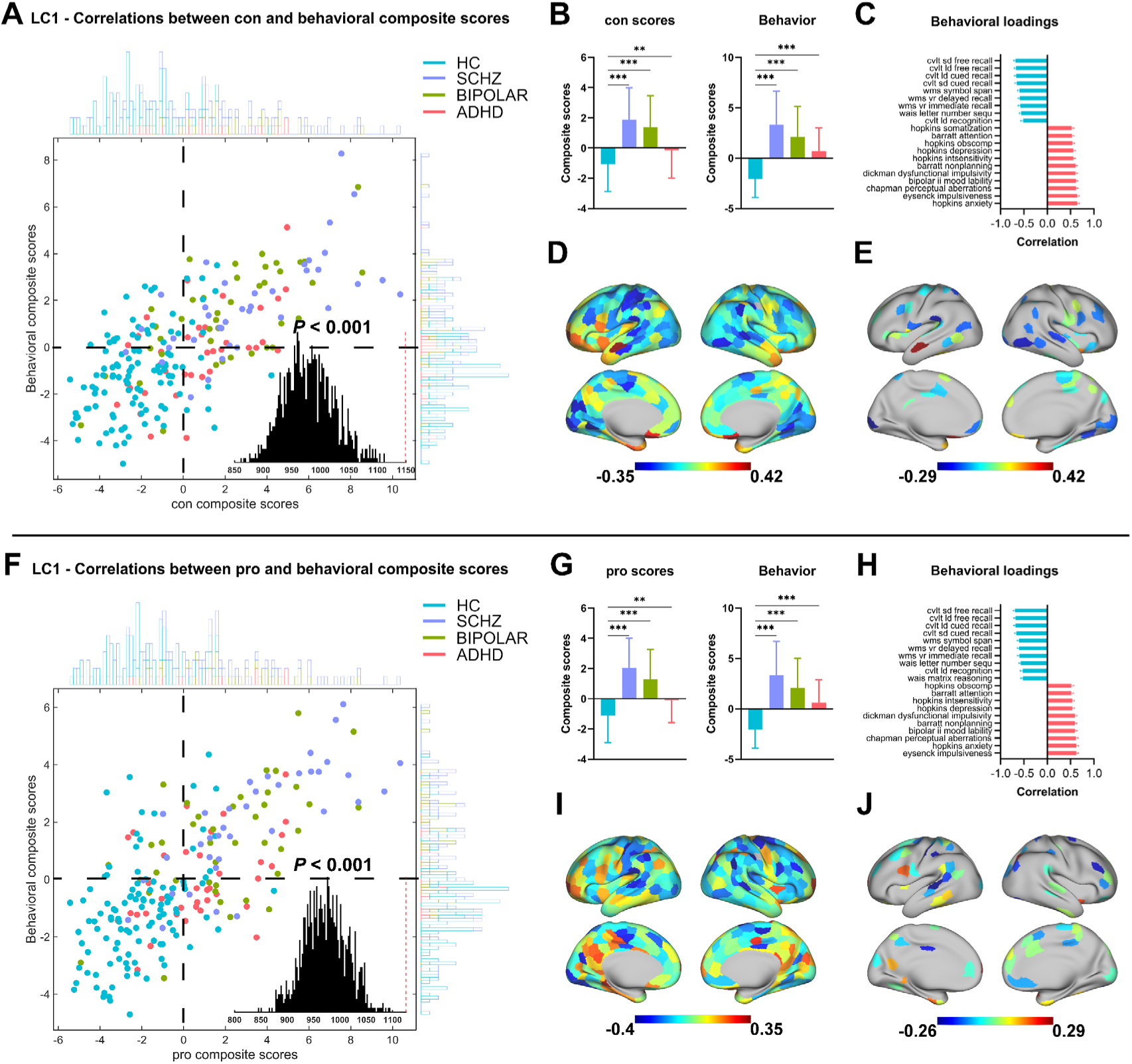
PLS analysis of UCLA dataset. Figures (A) to (E) show the results of LC1 for CON, and Figures (F) to (J) show the results of LC1 for PRO. (A) and (F) shows the correlation between the CON/PRO scores and participants’ behavioral composite scores. The inset illustrates the null distribution derived from permutation testing (1,000 iterations), with the actual singular value for LC1 indicated by a dashed line. Notably, the null distribution is not centered at zero. (B) and (G) shows the group differences in CON/PRO scores and behavioral composite scores. Note: Asterisks indicate t tests that survived false discovery rate correction (* q < .05; ** q <.01; *** q <.001). (C) and (H) shows the top 20 strongest Pearson correlations between participants’ behavioral indicators and their behavioral composite scores (CON/PRO). Error bars indicate bootstrapped standard deviations. (D) and (I) shows the unthresholded correlations between participants’ CON/PRO scores and their CON/PRO composite scores, with red indicating positive associations and blue indicating negative associations between PRO score and LC1. (E) and (J) shows the thresholded correlations between participants’ CON/PRO scores and the corresponding composite scores, only blocks with significant bootstrapped Z scores are shown (false discovery rate q < .05).

Comparisons across diagnostic groups revealed significant differences in LC1 hub composite scores and behavioral composite scores. Specifically, the HC group exhibited significantly lower LC1 hub composite scores and behavioral composite scores than the BD, ADHD, and SCHZ groups (Figures 3B and 3G). In the patient’s group comparison of **Table 2**, the hub composite scorers (CON and PRO) of LC1 in ADHD group significantly lower than SCHZ and BIPOLAR. Among the top 20 behavioral measures most correlated with the LC1 behavioral composite score, the only two differences between *CON* and *PRO* analyses were somatization symptoms and fluid intelligence, whereas the remaining measures were consistent across both hub types. These measures primarily reflect multidimensional assessments of psychological and cognitive function, emphasizing the broad cognitive and affective implications of hub properties. More specifically, LC1 behavioral composite scores were positively correlated with mental health and behavioral traits but negatively correlated with cognitive and working memory functions, indicating that higher hub connectivity in disease groups was linked to poorer cognitive performance but heightened emotional and behavioral reactivity.

**Table 2.** LC1 inter-group comparisons results of composite score.

| CON composite | p value | q value | ci1 | ci2 | t value | DF |
| --- | --- | --- | --- | --- | --- | --- |
| SCHZ-HC | 8.22675E-14 | 4.94E-13 | 2.244886 | 3.656696 | 8.261907 | 145 |
| BIPOLAR-HC | 7.02078E-11 | 2.11E-10 | 1.756857 | 3.130475 | 7.03104 | 148 |
| ADHD-HC | 0.008920247 | 0.010704 | 0.231494 | 1.588306 | 2.650892 | 145 |
| SCHZ-BIPOLAR | 0.293063643 | 0.293064 | -0.44697 | 1.461217 | 1.058856 | 75 |
| SCHZ-ADHD | 2.98614E-05 | 5.98E-05 | 1.128147 | 2.953636 | 4.457372 | 72 |
| BIPOLAR-ADHD | 0.001047072 | 0.001571 | 0.637891 | 2.429641 | 3.410541 | 75 |
| **behavior composite (CON based)** |  |  |  |  |  |  |
| SCHZ-HC | <0.00001 | <0.00001 | 4.538063 | 6.261019 | 12.38798 | 145 |
| BIPOLAR-HC | <0.00001 | <0.00001 | 3.411111 | 5.012565 | 10.39444 | 148 |
| ADHD-HC | 6.49392E-12 | 1.3E-11 | 2.038804 | 3.502782 | 7.48148 | 145 |
| SCHZ-BIPOLAR | 0.10738015 | 0.10738 | -0.26421 | 2.639611 | 1.629596 | 75 |
| SCHZ-ADHD | 0.000210254 | 0.000315 | 1.286805 | 3.970692 | 3.905018 | 72 |
| BIPOLAR-ADHD | 0.022769267 | 0.027323 | 0.20643 | 2.675662 | 2.325186 | 75 |
| **PRO composite** |  |  |  |  |  |  |
| SCHZ-HC | 5.55E-16 | 3.33E-15 | 2.476517 | 3.846566 | 9.121806 | 145 |
| BIPOLAR-HC | 4.59E-11 | 1.38E-10 | 1.733739 | 3.068429 | 7.110018 | 148 |
| ADHD-HC | 0.00192617 | 0.002311 | 0.384594 | 1.670047 | 3.15913 | 145 |
| SCHZ-BIPOLAR | 0.092329087 | 0.092329 | -0.12804 | 1.648955 | 1.705024 | 75 |
| SCHZ-ADHD | 1.22E-06 | 2.44E-06 | 1.331091 | 2.937349 | 5.297394 | 72 |
| BIPOLAR-ADHD | 0.000934176 | 0.001401 | 0.579698 | 2.167828 | 3.446413 | 75 |
| **behavior (PRO based)** |  |  |  |  |  |  |
| SCHZ-HC | <0.00001 | <0.00001 | 4.534184 | 6.267408 | 12.31746 | 145 |
| BIPOLAR-HC | <0.00001 | <0.00001 | 3.309068 | 4.911307 | 10.1386 | 148 |
| ADHD-HC | 3.35E-11 | 6.70E-11 | 1.937426 | 3.40918 | 7.180104 | 145 |
| SCHZ-BIPOLAR | 7.81E-02 | 0.0781 | -0.14891 | 2.730126 | 1.786035 | 75 |
| SCHZ-ADHD | 1.16E-04 | 1.74E-04 | 1.394303 | 4.060683 | 4.078308 | 72 |
| BIPOLAR-ADHD | 2.12E-02 | 0.02544 | 0.22054 | 2.653228 | 2.353298 | 75 |

Note: ci1: lower limit of confidence interval for differences in overall means; ci2: upper limit of confidence interval for differences in overall means; DF: Degrees of Freedom;

Network-level hub distributions further supported these findings. As shown in Figures 3D and 3E, higher CON composite scores were associated with increased CON scores in the limbic and DMN, whereas lower CON composite scores corresponded to increased CON scores in the visual VSN)and SMN. Similarly, as illustrated in Figures 3I and 3J, higher PRO composite scores were linked to increased *PRO* scores in the DMN and control networks, whereas lower *PRO* composite scores corresponded to increased *PRO* scores in the SMN network. These findings suggest that alterations in hub node distributions reflect network-specific reorganization patterns in different cognitive and clinical conditions. The top 20 ROIs with the strongest positive or negative correlations are detailed in **Supplementary Table S6**.

LC2 accounted for 14% of the covariance in CON scores and 15% in PRO scores, indicating a moderate contribution to hub-behavior relationships. As shown in Figures 4A and 4F, along with group comparison results in Figures 4B and 4G, the distributions and intergroup relationships between hub composite scores (CON and PRO) and behavioral composite scores exhibited distinct patterns.

**Figure 4.**
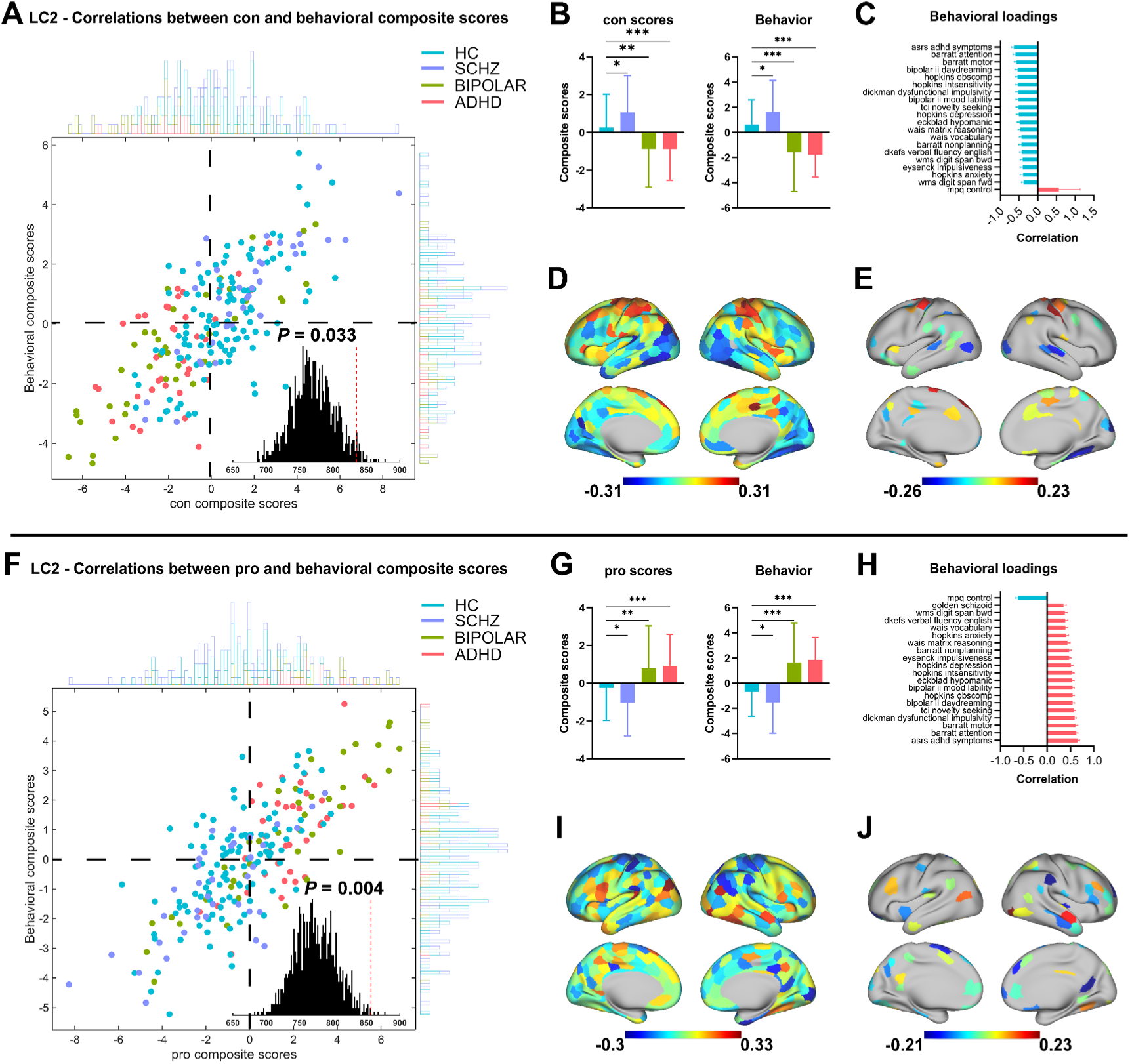
PLS analysis of UCLA dataset. Figures (A) to (E) show the results of LC2 for CON, and Figures (F) to (J) show the results of LC2 for PRO. (A) and (F) shows the correlation between the CON/PRO scores and participants’ behavioral composite scores. The inset illustrates the null distribution derived from permutation testing (1,000 iterations), with the actual singular value for LC1 indicated by a dashed line. Notably, the null distribution is not centered at zero. (B) and (G) shows the group differences in CON/PRO scores and behavioral composite scores. Note: Asterisks indicate t tests that survived false discovery rate correction (* q < .05; ** q < .01; *** q < .001). (C) and (H) shows the top 20 strongest Pearson correlations between participants’ behavioral indicators and their behavioral composite scores (CON/PRO). Error bars indicate bootstrapped standard deviations. (D) and (I) shows the unthresholded correlations between participants’ CON/PRO scores and their CON/PRO composite scores, with red indicating positive associations and blue indicating negative associations between PRO score and LC1. (E) and (J) shows the thresholded correlations between participants’ CON/PRO scores and the corresponding composite scores, only blocks with significant bootstrapped Z scores are shown (false discovery rate q < .05).

For CON scores, the HC group showed significantly lower composite scores than the SCHZ group but higher scores than the BIPOLAR and ADHD groups (Figure 4B). In contrast, for PRO scores, the HC group demonstrated an inverse pattern (Figure 4G), suggesting that different hub properties may be differentially affected across diagnostic groups. Among the patient groups comparison of **Table 3**, the SCHZ group exhibited significantly higher CON comprehensive scores and corresponding behavioral comprehensive scores compared to the BIPOLAR and ADHD groups. While the PRO comprehensive scores and their corresponding behavioral comprehensive scores were significantly lower in the SCHZ group than in the BIPOLAR and ADHD groups. Despite these variations, both CON and PRO composite scores remained significantly positively correlated with behavioral composite scores (*r_con_* = 0.696, *r_pro_* = 0.743, *p_(con/pro)_* < 0.001), reinforcing the functional relevance of hub network configurations.

**Table 3.** LC2 inter-group comparisons results of composite score.

| CON composite | p value | q value | ci1 | ci2 | t value | DF |
| --- | --- | --- | --- | --- | --- | --- |
| SCHZ-HC | 0.022676 | 0.027212 | 0.112822 | 1.476798 | 2.303428 | 145 |
| BIPOLAR-HC | 0.00134 | 0.002009 | -1.78515 | -0.44018 | -3.26959 | 148 |
| ADHD-HC | 0.000935 | 0.001871 | -1.77448 | -0.46466 | -3.37878 | 145 |
| SCHZ-BIPOLAR | 7.79E-05 | 0.000234 | 0.998532 | 2.816416 | 4.180558 | 75 |
| SCHZ-ADHD | 2.31E-05 | 0.000139 | 1.071502 | 2.75726 | 4.527635 | 72 |
| BIPOLAR-ADHD | 0.987139 | 0.987 | -0.84382 | 0.857638 | 0.016173 | 75 |
| **behavior composite (CON based)** |  |  |  |  |  |  |
| SCHZ-HC | 0.012295 | 0.014754 | 0.222793 | 1.798666 | 2.535314 | 145 |
| BIPOLAR-HC | 8.1E-07 | 1.62E-06 | -3.04958 | -1.35876 | -5.15218 | 148 |
| ADHD-HC | 7.91E-10 | 4.75E-09 | -3.1082 | -1.67267 | -6.58236 | 145 |
| SCHZ-BIPOLAR | 4.27E-06 | 6.41E-06 | 1.92367 | 4.506126 | 4.959934 | 75 |
| SCHZ-ADHD | 3.51E-09 | 1.05E-08 | 2.393216 | 4.409112 | 6.726634 | 72 |
| BIPOLAR-ADHD | 0.751967 | 0.752 | -0.9835 | 1.356032 | 0.317208 | 75 |
| **PRO composite** |  |  |  |  |  |  |
| SCHZ-HC | 0.018025 | 0.02163 | -1.44104 | -0.13716 | -2.39229 | 145 |
| BIPOLAR-HC | 0.002846 | 0.004269 | 0.36609 | 1.733073 | 3.034569 | 148 |
| ADHD-HC | 0.00046 | 0.000919 | 0.524323 | 1.8128 | 3.58503 | 145 |
| SCHZ-BIPOLAR | 0.000147 | 0.000441 | -2.75434 | -0.92302 | -4.00023 | 75 |
| SCHZ-ADHD | 5.50E-06 | 3.30E-05 | -2.75248 | -1.16284 | -4.90993 | 72 |
| BIPOLAR-ADHD | 0.793024 | 0.793024 | -1.01909 | 0.78113 | -0.26332 | 75 |
| **behavior (PRO based)** |  |  |  |  |  |  |
| SCHZ-HC | 0.037115 | 0.044538 | -1.60521 | -0.05013 | -2.10388 | 145 |
| BIPOLAR-HC | 2.28E-07 | 4.56E-07 | 1.481968 | 3.178626 | 5.42826 | 148 |
| ADHD-HC | 4.10E-11 | 2.46E-10 | 1.856103 | 3.276378 | 7.142377 | 145 |
| SCHZ-BIPOLAR | 6.39E-06 | 9.59E-06 | -4.45363 | -1.8623 | -4.85542 | 75 |
| SCHZ-ADHD | 2.18E-09 | 6.54E-09 | -4.38315 | -2.40467 | -6.83923 | 72 |
| BIPOLAR-ADHD | 6.92E-01 | 0.692 | -1.41706 | 0.945177 | -0.39795 | 75 |

Note: ci1: lower limit of confidence interval for differences in overall means; ci2: upper limit of confidence interval for differences in overall means; DF: Degrees of Freedom;

Among the top 20 behavioral measures most strongly correlated with behavioral composite scores, only two measures differed between CON and PRO analyses: working memory tests and schizotypal personality traits. The remaining measures were consistent and primarily reflected cognitive flexibility, abstract reasoning, and verbal expression, emphasizing the role of network hubs in higher-order cognitive processes. For CON, the LC2 behavioral composite score was negatively correlated with cognitive measures but positively correlated with self-regulation (Figure 4C). In contrast, for PRO, this pattern was entirely reversed, as shown in Figure 4H, suggesting that different hub types contribute to distinct cognitive and behavioral profiles.

Examining the relationship between hub scores and composite scores at the network level further supports this distinction. Higher CON composite scores were associated with increased CON scores in the SMN and DAN networks, whereas lower CON composite scores were linked to elevated CON scores in the VSN network. For PRO, higher composite scores corresponded to increased PRO scores in the DMN network, whereas lower composite scores were associated with elevated PRO scores in the SMN network. These findings indicate that variability in hub node distributions across networks is systematically linked to behavioral and cognitive profiles across different psychiatric conditions. The top 20 ROIs with the strongest positive or negative correlations are detailed in **Supplementary Table S7**.

## Discussion

In this study, we present the Multi-Index Entropy Hub Score (MIEHS) as a novel and systematic approach to quantifying brain network hub properties using multiple graph-theoretic indicators. Through extensive validation across benchmark networks, random network simulations, and resting-state fMRI datasets, our findings demonstrate the accuracy, stability, and clinical relevance of MIEHS in identifying connector and provincial hubs. The results provide crucial insights into the hierarchical organization of brain networks and the role of hub alterations in psychiatric disorders.

Current hub identification methods predominantly rely on single-metric or qualitative approaches—for instance, betweenness centrality ^38,39^, degree centrality ^40^, node efficiency ^41^, or shortest path length ^42^—which inadequately capture the complex structural and functional properties of network hubs. Unlike computational methods like PageRank ^6^, eigenvector centrality ^43^, and Katz centrality ^44^, which also provide quantitative assessments of hub properties, MIEHS capability to evaluate hierarchical hub attributes—connector (CON) and provincial (PRO) hubs. This hierarchical assessment is crucial for the inherently layered structure of the human brain system. By integrating six graph-theoretic measures with entropy-based weighting, MIEHS offers a more robust and quantitative framework for distinguishing between connector hubs, which facilitate inter-network communication, and provincial hubs, which support local information processing. Compared to MQGA, MIEHS exhibits superior sensitivity in identifying hub properties, particularly for provincial hubs, where often confounded by global network sparsity.

Our benchmark network validation highlights MIEHS’s ability to reliably classify hub nodes, as evidenced by its application to *Zachary’s Karate Club* network and a simplified 12-node model. The alignment between hub rankings and diffusion dynamics, as illustrated by the SSI model, further reinforces the validity of the hub score computation. Additionally, large-scale simulations involving 100 random networks demonstrate that MIEHS achieves a significantly higher correlation between hub scores and node labels than MQGA, especially when network sparsity exceeds 0.2. This suggests that MIEHS is well-suited for hub classification across various network densities, making it a more adaptable tool for real-world fMRI applications.

### Functional Network Architecture and Stability of Hub Nodes

The analysis of resting-state fMRI data derived from the MSC dataset corroborates the validity of MIEHS in elucidating significant hub structures within the human brain’s connectome. This investigation included a stability analysis conducted across varying network sparsity thresholds ranging from 0.01 to 0.5. The results revealed that hub scores demonstrated remarkable consistency throughout the different thresholds, with provincial hub scores showing slightly more stability than connector hub scores. This finding is particularly significant considering that connector hubs are characterized by their role in integrating information from a broader array of global network connections, which makes them more vulnerable to fluctuations in network sparsity ^45^. Supporting evidence illustrates that stability in hub scores correlates positively with intraclass correlation (ICC) values, suggesting that hubs scoring high are inherently more stable over time, thereby reinforcing the biological relevance of these network properties (Cole et al., 2015b).

The spatial distribution of high-scoring hubs aligns with prevailing neurocognitive network hierarchies, where it has been noted that connector hubs are largely situated within the attention network. This positioning underscores their critical function in integrating higher-order cognitive processes, which are pivotal for tasks requiring focused and sustained attention ^47,48^. Provincial hubs, on the other hand, are primarily located within the DMN, which plays a vital role in self-referential and introspective processing. The partitioning of hub roles, i.e. connector hubs facilitating information flow between distinct network modules and provincial hubs enhancing connectivity within a given module, illuminates the necessity of both types in maintaining coherent overall brain function ^47^. Gradient mapping techniques further corroborate these findings by illustrating that connector hubs cluster effectively at the interfaces between unimodal and transmodal cortical regions. This anatomical arrangement reflects their functional significance in bridging sensory and associative cortical areas, essential for integrative processing ^49^. Conversely, provincial hubs display a more diffuse distribution across both unimodal and transmodal gradients, suggesting their broader role in stabilizing local networks. Such observations support the idea that provincial hubs contribute to increased modularity and local connectivity, facilitating dedicated processing pathways, while connector hubs are instrumental for reducing modular segregation by enhancing between-module connectivity ^50,51^.

Our findings also provide novel insights into hub stability across different functional networks. The global random null model validation revealed that the DMN contained the highest number of significant ROIs, further reinforcing its role as a key network hub. However, when normalizing for network size, the Limbic network exhibited the highest significance rate, with all its ROIs showing significance in the connector hub null model validation. This suggests that while the DMN serves as a major integrative hub in large-scale networks, the Limbic system plays a disproportionately critical role in local information processing and network resilience.

### Clinical Implications of Hub Alterations

To evaluate the generalizability of MIEHS, we applied it to four diagnostic groups in the UCLA dataset, revealing significant changes in hub distributions across psychiatric conditions. Despite differences in functional connectivity patterns between the MSC and UCLA healthy control groups, hub scores remained significantly correlated across datasets, with provincial hub scores demonstrating greater stability than connector hub scores. This further supports the robustness of provincial hub properties and highlights the impact of global connectivity disruptions on connector hub stability.

PLS analysis revealed strong associations between hub scores and behavioral traits, with LC1 accounting for 26% of the covariance in CON scores and 25% in PRO scores. Across diagnostic groups, the HC group exhibited significantly lower LC1 hub composite scores than the clinical groups, suggesting that increased hub connectivity in psychiatric populations may reflect maladaptive reorganization. Notably, LC1 hub composite scores were positively correlated with mental health and behavioral traits but negatively correlated with cognitive and working memory functions, indicating that atypical hub organization may underlie cognitive dysfunction in clinical populations.

The network-level distributions of hub changes further elucidate these effects. Higher CON composite scores were linked to increased connectivity within the Limbic and DMN networks, whereas lower CON composite scores corresponded to increased hub strength in the VSN and SMN networks. Similarly, higher PRO composite scores were associated with the DMN and control networks, while lower PRO scores were linked to increased SMN connectivity. These findings suggest that psychiatric disorders may be characterized by excessive integration within associative networks at the expense of sensorimotor system balance^52,53^ potentially leading to cognitive and behavioral impairments.

Further supporting this interpretation, LC2 accounted for 14% and 15% of the covariance in CON and PRO scores, respectively, but showed an inverse pattern of hub alterations across groups. In contrast to LC1, the HC group exhibited lower CON composite scores than SCHZ but higher scores than BIPOLAR and ADHD, while the pattern was reversed for PRO scores. This suggested that different neuropsychiatric disorders are tied to distinct alterations in CON and PRO hubs, reflecting varying impacts on large-scale network integration. Behavioral correlations further reinforce this, as higher LC2 CON composite scores were negatively associated with cognitive flexibility and reasoning but positively associated with self-regulation, whereas higher PRO composite scores followed the opposite trend. At the network level, higher LC2 CON composite scores were linked with greater connectivity in the SMN and DAN networks, while lower scores were associated with increased connectivity in the VSN network. Similarly, higher PRO scores corresponded to greater connectivity in the DMN, whereas lower scores were linked to increased PRO strength in the SMN. These findings further highlight the role of hub reorganization in psychiatric disorders ^54^, particularly in the balance between sensorimotor and associative networks. The SMN network emerges as a critical player, with its hub property changes being a major contributor to variations across psychiatric disorders and LC1/LC2. Emerging evidence suggests that disruptions in the SMN may contribute to psychiatric disorders by impairing sensorimotor integration and motor control, potentially underlying the motor and cognitive symptoms observed in multiple mental diseases ^31,55^.

### Limitations and Future Directions

While MIEHS offers a more comprehensive framework for hub identification, several limitations must be acknowledged. First, although our validation spanned multiple datasets and network configurations, further cross-cohort replication is necessary to ensure robustness across diverse populations. Second, while we focused on resting-state functional connectivity, future studies should investigate task-based network dynamics to assess how changes in hub structure impact cognitive processing. Finally, although MIEHS enhances hub detection compared to traditional methods, integrating machine learning approaches could further refine its predictive capabilities and clinical applicability.

## Conclusion

This study introduces MIEHS as a novel and effective method for quantifying brain network hub properties, demonstrating its validity across various datasets and network structures. By integrating multiple-theoretic metrics and entropy-based weighting, MIEHS offers a robust framework for differentiating between connector hubs and provincial hubs, revealing their distinct functional and clinical significance. Our findings highlight the stability of hub properties in healthy brains, while also demonstrating network reorganization in psychiatric disorders, particularly in the interplay between sensorimotor and associative networks. These insights pave the way for future investigations into hub dynamics in health and disease, with potential applications in precision psychiatry and network-based interventions.

## Acknowledgements

This work received no external funding. No financial support was provided by any organization for the conduct of the research, the preparation of the manuscript, or the decision to publish.

## Author contributions

H.W. conceived and designed the study, performed all data analyses, developed the MIEHS methods, and wrote the first draft of the manuscript. J.X. provided the preprocessed MSC and UCLA data, and engaged in regular discussion to refine the analytical approach. B.K.-B. and E.A. assisted in verifying analysis pipelines and in discussion of intermediate results. B.K.-B. led the critical revision and editing of the manuscript, substantially improving its clarity, presentation, and scientific argumentation. E.A., B.B., S.F., B.Bi., and M.M. each contributed to manuscript review and provided constructive feedback on study design, data interpretation, and writing. B.Bi. and M.M. jointly supervised the project, provided strategic guidance, and oversaw experimental progress. All authors read and approved the submitted version of the manuscript.

## Data availability

Midnight Scan Club (MSC) dataset

The resting–state fMRI data from the Midnight Scan Club are publicly available at the OpenNeuro repository under accession number ds000224, version 1.0.4: https://openneuro.org/datasets/ds000224/versions/1.0.4.

UCLA Consortium for Neuropsychiatric Phenomics (CNP) dataset

All data from the UCLA cohort were originally shared via the OpenfMRI database under accession number ds000030 and can be accessed here: https://openfmri.org/dataset/ds000030/.

---

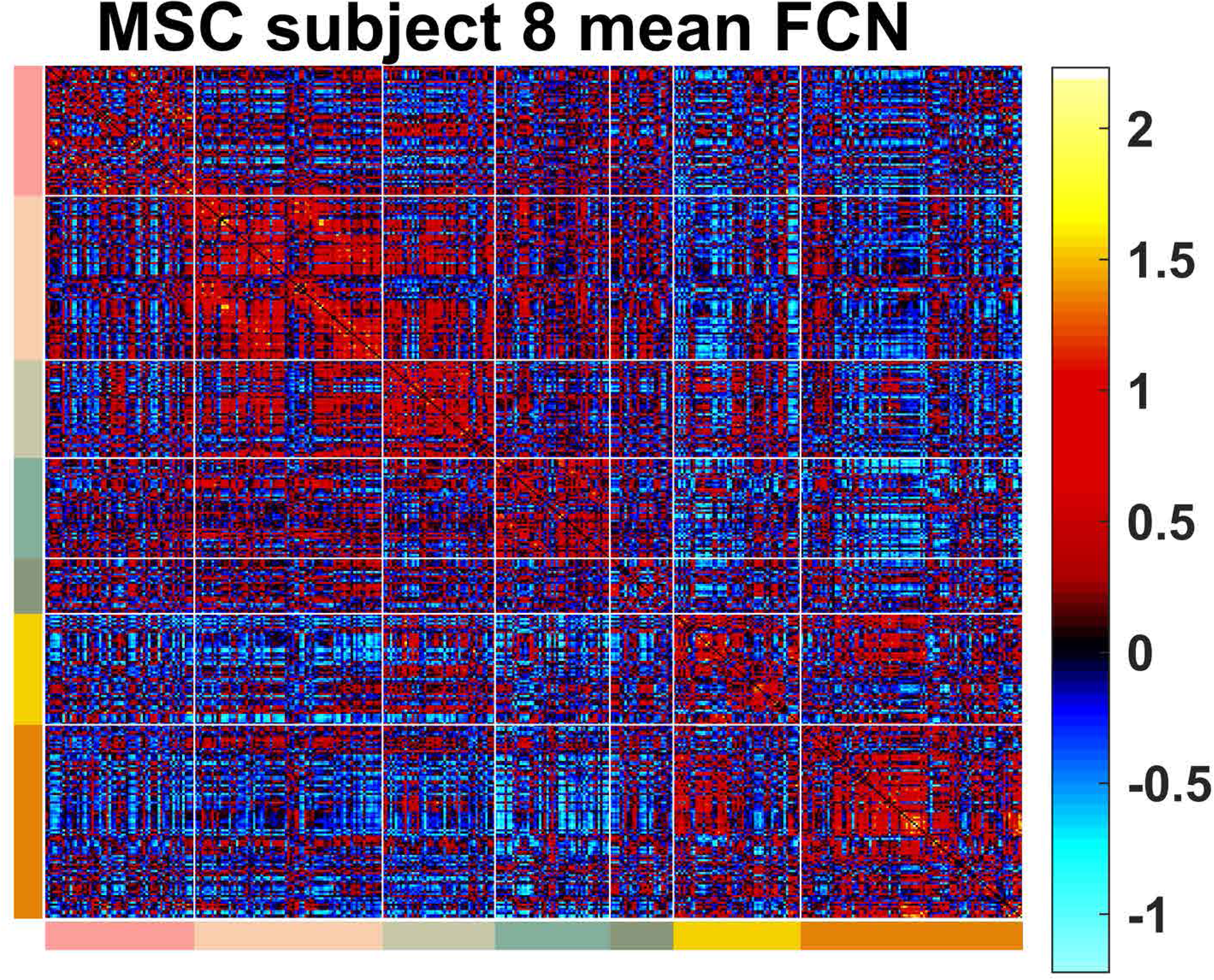

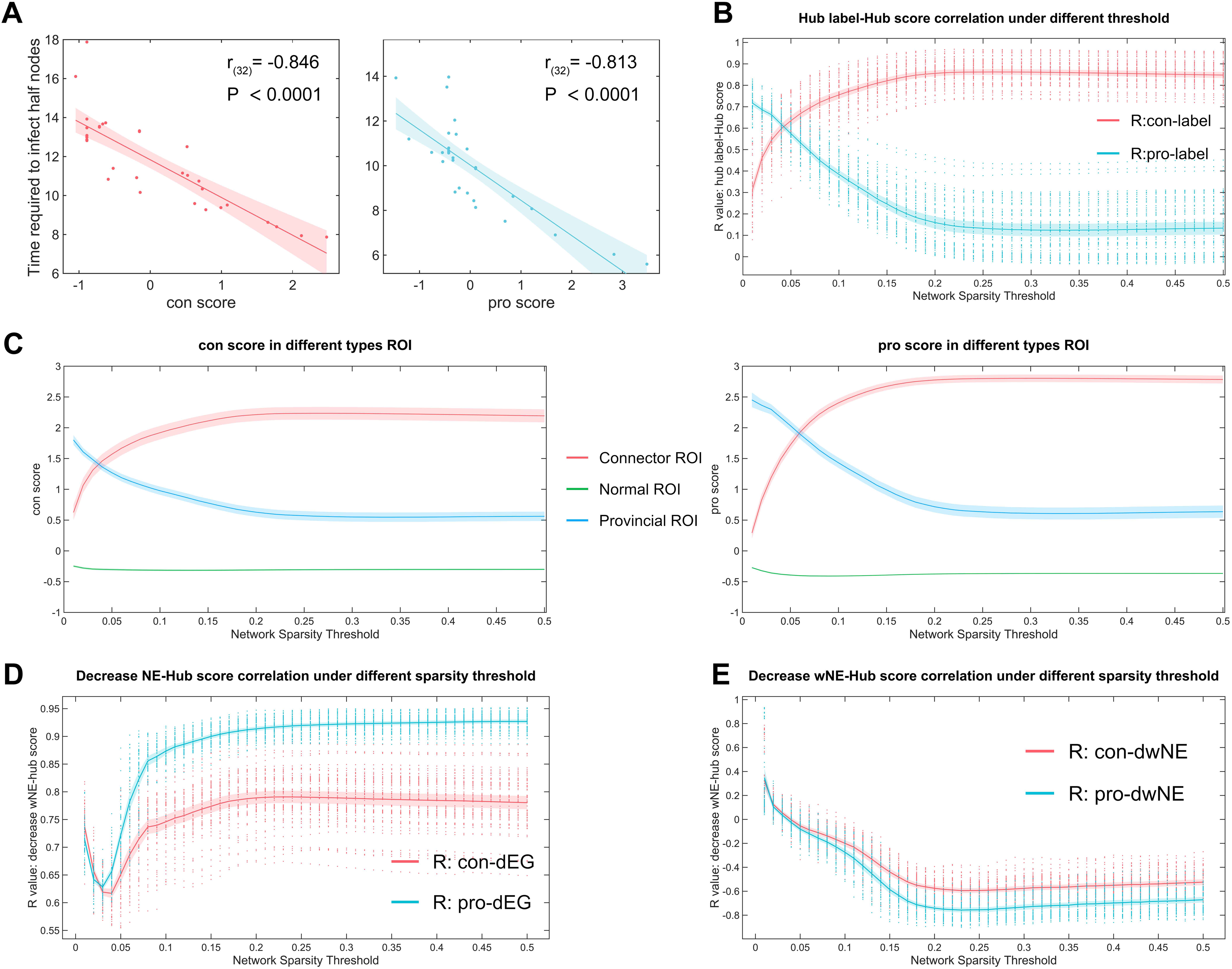

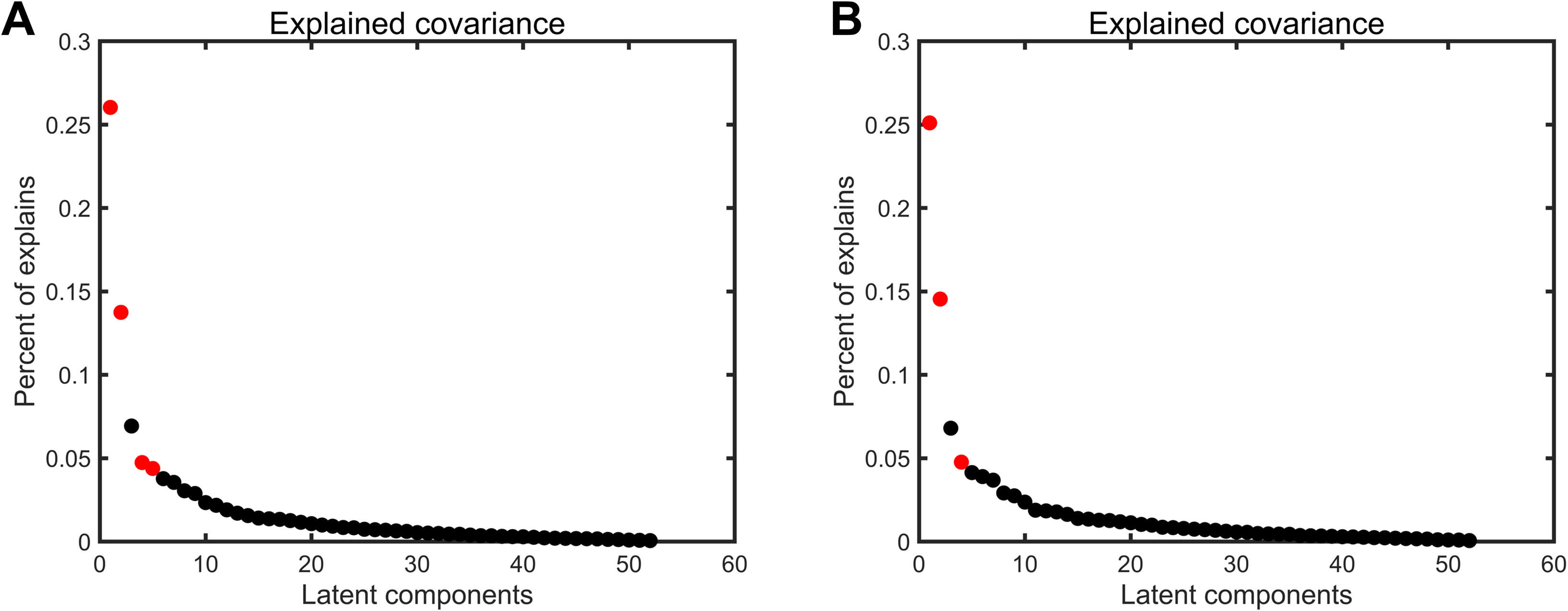

## References

1. Gu, S. et al. Controllability of structural brain networks. Nat Commun 6, (2015).

2. Bassett, D. S. & Bullmore, E. T. Small-World Brain Networks Revisited. Neuroscientist vol. 23 499–516 Preprint at 10.1177/1073858416667720 (2017).

3. Lord, L. D., Stevner, A. B., Deco, G. & Kringelbach, M. L. Understanding principles of integration and segregation using whole-brain computational connectomics: Implications for neuropsychiatric disorders. Philosophical Transactions of the Royal Society A: Mathematical, Physical and Engineering Sciences vol. 375 Preprint at 10.1098/rsta.2016.0283 (2017).

4. Fair, D. A. et al. Development of distinct control networks through segregation and integration. Proc Natl Acad Sci U S A 104, 13507–13512 (2007).

5. Finc, K. et al. Dynamic reconfiguration of functional brain networks during working memory training. Nat Commun 11, (2020).

6. Power, J. D., Schlaggar, B. L., Lessov-Schlaggar, C. N. & Petersen, S. E. Evidence for hubs in human functional brain networks. Neuron 79, 798–813 (2013).

7. Guimerà, R. & Amaral, L. A. N. Cartography of complex networks: Modules and universal roles. Journal of Statistical Mechanics: Theory and Experiment 2005, 1–13 (2005).

8. Bagarinao, E. et al. Connectivity impairment of cerebellar and sensorimotor connector hubs in Parkinson’s disease. Brain Commun 4, (2022).

9. Rotem-Kohavi, N. et al. Hub distribution of the brain functional networks of newborns prenatally exposed to maternal depression and SSRI antidepressants. Depress Anxiety 36, 753–765 (2019).

10. Yang, D. et al. Joint hub identification for brain networks by multivariate graph inference. Med Image Anal 73, (2021).

11. Korponay, C., Janes, A. C. & Frederick, B. B. Brain-wide functional connectivity artifactually inflates throughout functional magnetic resonance imaging scans. Nat Hum Behav 8, 1568–1580 (2024).

12. Teeuw, J., Hulshoff Pol, H. E., Boomsma, D. I. & Brouwer, R. M. Reliability modelling of resting-state functional connectivity. Neuroimage 231, (2021).

13. Bertolero, M. A., Yeo, B. T. T., Bassett, D. S. & D’Esposito, M. A mechanistic model of connector hubs, modularity and cognition. Nature Human Behaviour vol. 2 765–777 Preprint at 10.1038/s41562-018-0420-6 (2018).

14. Hwang, K., Bertolero, M. A., Liu, W. B. & D’Esposito, M. The human thalamus is an integrative hub for functional brain networks. Journal of Neuroscience 37, 5594–5607 (2017).

15. Ma, X. et al. Aberrant functional connectome in neurologically asymptomatic patients with end-stage renal disease. PLoS One 10, (2015).

16. Buckner, R. L. et al. Cortical hubs revealed by intrinsic functional connectivity: Mapping, assessment of stability, and relation to Alzheimer’s disease. Journal of Neuroscience 29, 1860–1873 (2009).

17. van den Heuvel, M. P. & Sporns, O. Network hubs in the human brain. Trends Cogn Sci 17, 683–696 (2013).

18. Crossley, N. A. et al. Cognitive relevance of the community structure of the human brain functional coactivation network. Proc Natl Acad Sci U S A 110, 11583–11588 (2013).

19. Li, Y. et al. Impaired Topological Properties of Gray Matter Structural Covariance Network in Epilepsy Children With Generalized Tonic–Clonic Seizures: A Graph Theoretical Analysis. Front Neurol 11, (2020).

20. Guo, Y. et al. Topological Alterations in White Matter Structural Networks in Blepharospasm. Movement Disorders 36, 2802–2810 (2021).

21. Liang, X., Zou, Q., He, Y. & Yang, Y. Coupling of functional connectivity and regional cerebral blood flow reveals a physiological basis for network hubs of the human brain. Proc Natl Acad Sci U S A 110, 1929–1934 (2013).

22. Cole, M. W., Ito, T. & Braver, T. S. Lateral prefrontal cortex contributes to fluid intelligence through multinetwork connectivity. Brain Connect 5, 497–504 (2015).

23. Cole, M. W., Pathak, S. & Schneider, W. Identifying the brain’s most globally connected regions. Neuroimage 49, 3132–3148 (2010).

24. Wu, H. et al. MQGA: A quantitative analysis of brain network hubs using multi-graph theoretical indices. Neuroimage 303, (2024).

25. Wu, Q. et al. Test-Retest Reliability of Resting Brain Small-World Network Properties across Different Data Processing and Modeling Strategies. Brain Sci 13, (2023).

26. Girvan, M. & Newman, M. E. J. Community structure in social and biological networks. Proc Natl Acad Sci U S A 99, 7821–7826 (2002).

27. Pastor-Satorras, R. & Vespignani, A. Epidemic spreading in scale-free networks. Phys Rev Lett 86, 3200–3203 (2001).

28. Gordon, E. M. et al. Precision Functional Mapping of Individual Human Brains. Neuron 95, 791–807.e7 (2017).

29. Poldrack, R. A. & Gorgolewski, K. J. OpenfMRI: Open sharing of task fMRI data. Neuroimage 144, 259–261 (2017).

30. Poldrack, R. A. et al. A phenome-wide examination of neural and cognitive function. Sci Data 3, (2016).

31. Kebets, V. et al. Somatosensory-Motor Dysconnectivity Spans Multiple Transdiagnostic Dimensions of Psychopathology. Biol Psychiatry 86, 779–791 (2019).

32. Thomas Yeo, B. T., et al. The organization of the human cerebral cortex estimated by intrinsic functional connectivity. J Neurophysiol 106, 1125–1165 (2011).

33. Schaefer, A. et al. Local-global parcellation of the human cerebral cortex from intrinsic functional connectivity mri. Cerebral Cortex 28, 3095–3114 (2018).

34. Margulies, D. S. et al. Situating the default-mode network along a principal gradient of macroscale cortical organization. Proc Natl Acad Sci U S A 113, 12574–12579 (2016).

35. Markello, R. D. et al. neuromaps: structural and functional interpretation of brain maps. Nat Methods 19, 1472–1479 (2022).

36. McIntosh, A. R. & Lobaugh, N. J. Partial least squares analysis of neuroimaging data: Applications and advances. in NeuroImage vol. 23 (2004).

37. McIntosh, A. R. & Mišić, B. Multivariate statistical analyses for neuroimaging data. Annual Review of Psychology vol. 64 499–525 Preprint at 10.1146/annurev-psych-113011-143804 (2013).

38. van den Heuvel, M. P., Scholtens, L. H. & de Reus, M. A. Topological organization of connectivity strength in the rat connectome. Brain Struct Funct 221, 1719–1736 (2016).

39. Bullmore, E. & Sporns, O. The economy of brain network organization. Nature Reviews Neuroscience vol. 13 336–349 Preprint at 10.1038/nrn3214 (2012).

40. Harriger, L., van den Heuvel, M. P. & Sporns, O. Rich club organization of macaque cerebral cortex and its role in network communication. PLoS One 7, (2012).

41. Feng, M. et al. Decreased Local Specialization of Brain Structural Networks Associated with Cognitive Dysfuntion Revealed by Probabilistic Diffusion Tractography for Different Cerebral Small Vessel Disease Burdens. Mol Neurobiol 61, 326–339 (2024).

42. Chen, J. et al. Variation in Brain Subcortical Network Topology Between Men With and Without PE: A Diffusion Tensor Imaging Study. Journal of Sexual Medicine 17, 48–59 (2020).

43. Stam, C. J. Modern network science of neurological disorders. Nature Reviews Neuroscience vol. 15 683–695 Preprint at 10.1038/nrn3801 (2014).

44. Bonacich, P. & Lloyd, P. Eigenvector-like measures of centrality for asymmetric relations. Soc Networks 23, 191–201 (2001).

45. Hwang, K., Bertolero, M. A., Liu, W. B. & D’Esposito, M. The human thalamus is an integrative hub for functional brain networks. Journal of Neuroscience 37, 5594–5607 (2017).

46. Cole, M. W., Ito, T. & Braver, T. S. Lateral Prefrontal Cortex Contributes to Fluid Intelligence Through Multinetwork Connectivity. Brain Connect 5, 497–504 (2015).

47. Bagarinao, E. et al. Connectivity impairment of cerebellar and sensorimotor connector hubs in Parkinson’s disease. Brain Commun 4, (2022).

48. Zhou, Y. et al. The Hierarchical Organization of the Default, Dorsal Attention and Salience Networks in Adolescents and Young Adults. Cerebral Cortex 28, (2018).

49. Spadone, S. et al. Dynamic reorganization of human resting-state networks during visuospatial attention. Proc Natl Acad Sci U S A 112, 8112–8117 (2015).

50. Wang, B. et al. Network enhancement as a general method to denoise weighted biological networks. Nat Commun 9, (2018).

51. Bertolero, M. A., Thomas Yeo, B. T. & D’Esposito, M. The modular and integrative functional architecture of the human brain. Proc Natl Acad Sci U S A 112, E6798–E6807 (2015).

52. Bi, B., Che, D. & Bai, Y. Neural network of bipolar disorder: Toward integration of neuroimaging and neurocircuit-based treatment strategies. Translational Psychiatry vol. 12 Preprint at 10.1038/s41398-022-01917-x (2022).

53. Rungratsameetaweemana, N. Understanding Motor Abnormalities in Psychiatric Disorders as Altered Sensorimotor Processing. Biological Psychiatry Global Open Science vol. 1 Preprint at 10.1016/j.bpsgos.2021.06.006 (2021).

54. Mi, T. M. et al. Altered Functional Segregated Sensorimotor, Associative, and Limbic Cortical-Striatal Connections in Parkinson’s Disease: An fMRI Investigation. Front Neurol 12, (2021).

55. Kaiser, R. H., Andrews-Hanna, J. R., Wager, T. D. & Pizzagalli, D. A. Large-scale network dysfunction in major depressive disorder: A meta-analysis of resting-state functional connectivity. JAMA Psychiatry 72, 603–611 (2015).

