## Supplement Materials for "MIEHS: A Quantitative Approach to Hub Property Analysis in Brain Networks"

*Supplemental Information*

### Supplemental Methods

#### Graph Theory Index Calculation

Degree Centrality (DC):

$$DC_i = \sum_{j=1}^{n} w_{ij} \tag{1}$$

$DC_i$ represent the weighted degree centrality of node *i*, and $w_{ij}$ is the weight of the edge between node *i* and node *j*.

Betweenness Centrality (BC):

$$BC_i = \sum_{s \neq i \neq t} \frac{n_{st}^i}{g_{st}} \tag{2}$$

$BC_i$ represent the weighted betweenness centrality of node *i*, $n_{st}^i$ represent the number of shortest paths between nodes *s* and *t* passing through node *i*. $g_{st}$ represents the total number of shortest paths between nodes *s* and *t*.

Participation Coefficient (PC):

$$PC_i = 1 - \sum_{s=1}^{M} \left(\frac{k_{is}}{k_i}\right)^2 \tag{3}$$

$PC_i$ represent the weighted participation coefficient of node *i*, $k_{is}$ represents the sum of the weights of the edges connecting node i and nodes in community s, $k_i$ represents the sum of the weights of the edges connecting node *i* to all nodes, and *M* is the total number of communities.

Clustering Coefficient (CC):

$$\mathrm{CC}_i = \frac{1}{s_i(k_i - 1)} \sum_{j,k} \frac{w_{ij} + w_{ik}}{2} a_{ij} a_{ik} a_{jk} \tag{4}$$

$\mathrm{CC}_i$ represents the weighted clustering coefficient of node *i*, $s_i$ represents the weighted degree centrality of node *i*, $k_i$ represents the degree centrality of node *i*, $w_{ij}$ and $w_{ik}$ are the weights of the edges between node *i* and node *j* and node *k* respectively. $a_{ij}$, $a_{ik}$, $a_{jk}$ are the connection relationships between node *i* and node *j*, node *i* and node *k*, node *j* and node *k* respectively (1 if there is an edge, otherwise 0).

within-module betweenness centrality (wBC):

$$wBC_i = \sum_{s \neq i \neq t \in M} \frac{n_{st}^i}{g_{st}} \tag{5.1}$$

$$Z_{wBC_i} = \frac{wBC_i - \mu_{wBC_i}}{\sigma_{wBC_i}} \tag{5.2}$$

within-module degree centrality (wDC):

$$wDC_i = \sum_{i \neq j \in M} w_{ij} \tag{6.1}$$

$$Z_{wDC_i} = \frac{wDC_i - \mu_{wDC_i}}{\sigma_{wDC_i}} \tag{6.2}$$

The calculation method of wBC and wDC is similar to BC and DC, except that only BC (or DC) values within different modules are calculated. After the calculation is completed, we perform Z-standardization on it in order to analyze the relative characteristics of the nodes and make comparisons across modules.

#### Simplified Susceptible-Infected (SSI) model

The SSI model used in this study is based on the infection criterion of the original SI model, but in order to simplify the calculation, the recovery of infected nodes is not considered. The model operates with the following parameters: 1) **Raw network:** The '*Zachary*' network mentioned in the text. 2) **Transmission Rate (TR):** The probability of infection transmission per time step, the transmission rate in this study was set to 0.1. 3) **Number of Simulations:** Independent run 5,000 times to ensure robustness of the results. 4) **Simulation Time Steps:** The maximum temporal duration for each simulation. 5) **Initial Infected Nodes:** For each simulated infection process, we used 34 nodes as the initial infection sources separately.

The infection process was modeled iteratively for each time step *t*: 1) At time *t=0*, except for the initial infection point (*I(0)*), all nodes in the network are set to be uninfected. 2) For each infected node i, its neighbors j have a probability TR of becoming infected in the next step. 3) The total number of infected nodes *I(t)* at each time step is recorded.

To enhance the reliability of results, this process was repeated $N_{sim}$ times, yielding $I_t^{(sim)}$, the number of infected nodes at each time step for simulation *sim*. The mean infection curve across simulations, $\overline{I}(t)$, was computed as:

$$\overline{I}(t) = \frac{1}{N_{sim}} \sum_{sim=1}^{N_{sim}} I_t^{(sim)}(t) \tag{7}$$

#### MSC MRI data acquisition

The study was approved by the Washington University School of Medicine Human Studies Committee and Institutional Review Board, with informed consent obtained from all participants. Imaging for each subject was performed on a Siemens TRIO 3T

MRI scanner over the course of 12 sessions conducted on separate days, each beginning at midnight. Structural MRI was conducted across two separate days. In total, four T1-weighted images (sagittal, 224 slices, 0.8 mm isotropic resolution, TE=3.74 ms, TR=2400 ms, TI=1000 ms, flip angle = 8 degrees), four T2-weighted images (sagittal, 224 slices, 0.8 mm isotropic resolution, TE=479 ms, TR=3200 ms), four MRA (transverse, 0.6 × 0.6, × 1.0mm, 44 slices, TR=25ms, TE=3.34ms) and eight MRVs, including four in coronal and four in sagittal orientations (sagittal: 0.8 × 0.8 × 2.0mm thickness, 120 slices, TR=27ms, TE=7.05ms; coronal: 0.7 × 0.7 × 2.5mm thickness, 128 slices, TR=28ms TE= 7.18ms), were obtained for each subject. Analyses of the MRA and MRV scans are not reported here.

Referring to the research of *Gordon EM* et al. [1], we process the resting state fMRI data by the same. The main preprocessing steps included: remove first 5 time points, demeaning, detrending, regression of whole brain, ventricular, and white matter signals, and interpolation across censored frames to allow continuous band-pass filtering (0.009 Hz < f < 0.08 Hz). Censored frames were then removed from subsequent analyses. Surface processing of the data was conducted by sampling the BOLD fMRI volumetric timeseries onto each subject's mid-thickness cortical surfaces using Connectome Workbench's ribbon-constrained sampling. Based on the Yeo7 400ROIs template, the sampled data were resampled onto the standardized 32k fs_LR surface, facilitating direct comparisons across subjects [2]. Lastly, resting-state time courses were smoothed with 2 mm Gaussian kernels, see the original for more details [1].

#### UCLA MRI data acquisition

All participants provided written informed consent following a verbal explanation of the study, in line with Institutional Review Board approvals from UCLA and the Los Angeles County Department of Mental Health. All structural and functional UCLA MRI data were collected on two 3T Siemens Trio scanners. High-resolution anatomical data were collected by Magnetization Prepared Rapid Acquisition Gradient Echo (MPRAGE) scans using following parameters: TR=1.9 s, TE=2.26 ms, FOV =250 mm, matrix =256 × 256, sagittal plane, slice thickness=1 mm, 176 slices. Functional data were collected using a T2*-weighted echoplanar imaging sequence (EPI) acquired using following parameters: slice thickness=4 mm, 34 slices, TR=2 s, TE=30 ms, flip angle=90°, matrix 64 × 64, FOV=192 mm, oblique slice orientation.

All resting-state fMRI data were preprocessed using the fMRIPrep pipeline [3], with the following steps applied to each participant: removal of the first five time points, skull stripping, motion correction, slice timing correction, susceptibility distortion correction, and co-registration of functional and anatomical scans. Motion artifacts were automatically removed from the preprocessed BOLD time series via independent component analysis (ICA-AROMA), where ICA-AROMA motion components were used as noise regressors alongside mean physiological signals from white matter and

CSF. The time series were band-pass filtered between 0.009 Hz and 0.08 Hz.

#### Supplemental Results

##### Supplementary Tables

Table S1. The score of hub nodes in the Z network and the time required to infect half of the nodes.

| ROI | CON score | CON: Half I(t) | PRO score | PRO: Half I(t) |
|---|---|---|---|---|
| 1 | 2.005 | 7.94 | 2.764 | 5.597 |
| 2 | 0.82 | 9.269 | 1.05 | 7.515 |
| 3 | 1.843 | 8.397 | 0.174 | 8.075 |
| 4 | -0.057 | 10.16 | 0.42 | 8.133 |
| 5 | -0.666 | 13.509 | -0.441 | 10.354 |
| 6 | -0.128 | 13.319 | 0.243 | 9.867 |
| 7 | -0.128 | 13.281 | 0.243 | 9.917 |
| 8 | -0.503 | 10.833 | 0.111 | 8.822 |
| 9 | 1.045 | 9.378 | -0.761 | 10.748 |
| 10 | 0.437 | 12.505 | -1.617 | 13.929 |
| 11 | -0.666 | 13.545 | -0.441 | 10.245 |
| 12 | -1.238 | 16.108 | -1.557 | 13.518 |
| 13 | -0.917 | 13.472 | -0.095 | 10.791 |
| 14 | 0.676 | 9.587 | -0.326 | 8.776 |
| 15 | -0.917 | 12.817 | -0.104 | 10.589 |
| 16 | -0.917 | 12.845 | -0.104 | 10.601 |
| 17 | -0.917 | 17.876 | -0.095 | 13.968 |
| 18 | -0.917 | 12.994 | -0.095 | 10.589 |
| 19 | -0.917 | 12.872 | -0.104 | 10.66 |
| 20 | 0.671 | 10.741 | -1.022 | 10.588 |
| 21 | -0.917 | 12.817 | -0.104 | 10.605 |
| 22 | -0.917 | 13.082 | -0.095 | 10.569 |
| 23 | -0.917 | 12.842 | -0.104 | 10.671 |
| 24 | -0.122 | 10.916 | 0.769 | 8.434 |
| 25 | -0.632 | 13.672 | -0.851 | 12.039 |
| 26 | -0.601 | 13.729 | -0.851 | 11.408 |
| 27 | -0.917 | 13.923 | -0.104 | 11.457 |
| 28 | 0.535 | 11.034 | -0.906 | 10.186 |
| 29 | 0.407 | 11.151 | -1.014 | 11.195 |
| 30 | -0.442 | 11.395 | 0.032 | 9.002 |
| 31 | 0.729 | 10.336 | -0.76 | 10.595 |
| 32 | 1.186 | 9.512 | 1.137 | 8.625 |
| 33 | 1.638 | 8.623 | 1.917 | 6.903 |

| | | | | |
|---|---|---|---|---|
| 34 | 2.358 | 7.868 | 2.689 | 6.034 |

Note: Infecting half of the nodes in PRO refers to the time required to infect half of the nodes in the sub-network, which is different from the time required to infect the global network in CON.

Table S2. Basic information of subjects in the MSC dataset.

| Subjects | Gender | Age | Education (years) |
|---|---|---|---|
| 1 | Male | 34 | 22 |
| 2 | Male | 34 | 28 |
| 3 | Female | 29 | 18 |
| 4 | Female | 28 | 22 |
| 5 | M | 27 | 20 |
| 6 | Female | 24 | 17.5 |
| 7 | Female | 31 | 20 |
| 8 | Female | 27 | 21 |
| 9 | Male | 26 | 19 |
| 10 | Male | 31 | 19 |
| **AVG** | – | **29.1** | **20.7** |
| **SD** | – | **3.3** | **3** |

Table S3. Basic information of subjects in the UCLA dataset.

| Group | Male/Female | Age, mean (SD) | Education, mean (SD) |
|---|---|---|---|
| HC (n = 110) | 57/53 | 31.26 (8.66) | 15.19 (1.59) |
| ADHD (n = 37) | 20/17 | 31.22 (10.11) | 14.57 (1.85) |
| BD (n = 40) | 23/17 | 34.78 (9.24) | 14.43 (1.91) |
| SCHZ (n = 37) | 29/8 | 35.54 (9.08) | 12.78 (1.55) |

Table S4. Top 10% hub scoring nodes in the MSC dataset.

| ROI Name | mean con | ROI Name | mean pro |
|---|---|---|---|
| RH_SalVentAttn_TempOccPar_6 | 1.8190545 | LH_Default_Par_4 | 1.9689722 |
| LH_SalVentAttn_ParOper_4 | 1.451333 | LH_Default_PFC_13 | 1.6764686 |
| RH_SalVentAttn_Med_1 | 1.4274379 | RH_Default_pCunPCC_4 | 1.5724914 |
| RH_SalVentAttn_TempOccPar_7 | 1.3625436 | RH_Cont_Par_6 | 1.5465702 |
| LH_SalVentAttn_FrOperIns_9 | 1.3230781 | RH_Default_PFCdPFCm_4 | 1.5234562 |
| RH_DorsAttn_Post_14 | 1.2011474 | LH_Cont_Par_5 | 1.4388324 |
| RH_SalVentAttn_FrOperIns_2 | 1.1983706 | LH_Default_pCunPCC_6 | 1.4222909 |
| RH_SomMot_22 | 1.1884956 | LH_Default_pCunPCC_7 | 1.3644254 |
| RH_SalVentAttn_Med_6 | 1.1873091 | LH_Default_Par_6 | 1.3062506 |
| LH_SalVentAttn_Med_1 | 1.1751777 | RH_Default_PFCdPFCm_7 | 1.3035232 |
| RH_SalVentAttn_FrOperIns_5 | 1.1744858 | RH_SalVentAttn_Med_1 | 1.2867815 |

| | | | |
|---|---|---|---|
| RH_SalVentAttn_FrOperIns_7 | 1.1253284 | RH_Cont_Par_2 | 1.2398211 |
| RH_SalVentAttn_FrOperIns_8 | 1.1058161 | LH_DorsAttn_Post_12 | 1.2329236 |
| RH_SalVentAttn_Med_3 | 1.0963801 | RH_Cont_PFCl_5 | 1.2138785 |
| LH_Cont_Par_1 | 1.0638813 | RH_Cont_PFCl_9 | 1.2048009 |
| RH_DorsAttn_Post_11 | 1.062922 | LH_Cont_Par_4 | 1.203357 |
| RH_Cont_Par_2 | 1.0537237 | RH_Default_Par_3 | 1.1978693 |
| RH_SalVentAttn_TempOccPar_5 | 1.0252414 | RH_Default_pCunPCC_8 | 1.1770765 |
| LH_SalVentAttn_Med_4 | 1.0182133 | RH_Vis_6 | 1.1769484 |
| RH_SalVentAttn_TempOccPar_3 | 1.013901 | LH_DorsAttn_Post_9 | 1.1582487 |
| LH_Vis_30 | 1.0072826 | RH_DorsAttn_Post_12 | 1.1340288 |
| RH_Cont_Par_4 | 1.0044737 | RH_Vis_26 | 1.1112107 |
| LH_DorsAttn_Post_6 | 0.9823256 | LH_Cont_Par_6 | 1.1070656 |
| LH_Cont_Par_6 | 0.9785006 | LH_Default_PFC_19 | 1.1059952 |
| RH_Cont_pCun_1 | 0.9756555 | RH_SomMot_12 | 1.0858079 |
| RH_Default_pCunPCC_1 | 0.961386 | RH_Vis_5 | 1.0760109 |
| LH_SalVentAttn_FrOperIns_2 | 0.9316672 | LH_Vis_5 | 1.0719113 |
| LH_SalVentAttn_FrOperIns_7 | 0.9295387 | RH_SalVentAttn_TempOccPar_6 | 1.0574946 |
| LH_Vis_31 | 0.914214 | RH_Default_pCunPCC_5 | 1.0573341 |
| RH_DorsAttn_Post_13 | 0.8690651 | LH_Vis_4 | 1.0278897 |
| LH_DorsAttn_Post_9 | 0.855515 | LH_Default_PFC_14 | 1.0210837 |
| LH_SalVentAttn_ParOper_3 | 0.821724 | RH_DorsAttn_Post_14 | 1.0138913 |
| LH_DorsAttn_PrCv_1 | 0.8188136 | LH_Default_Par_2 | 0.9951074 |
| RH_SomMot_23 | 0.813755 | LH_Default_PFC_9 | 0.9815267 |
| RH_SalVentAttn_Med_4 | 0.811859 | RH_Vis_19 | 0.9749509 |
| LH_SalVentAttn_PFCl_1 | 0.8061318 | LH_DorsAttn_Post_7 | 0.9630879 |
| LH_SalVentAttn_ParOper_1 | 0.8054345 | RH_Cont_Par_5 | 0.9612197 |
| LH_Vis_29 | 0.8024193 | LH_Default_pCunPCC_11 | 0.9593565 |
| RH_Cont_PFCl_5 | 0.7841973 | RH_SomMot_10 | 0.9408174 |
| LH_DorsAttn_Post_12 | 0.7784574 | RH_SomMot_14 | 0.9174919 |

Table S5. Nodes significantly higher than the random null model and their corresponding Z-scores.

| **ROI Name** | **con: Z** | **ROI Name** | **pro: Z** |
|---|---|---|---|
| RH_Cont_pCun_1 | 47.205 | RH_Cont_pCun_1 | 12.95 |
| RH_Cont_Par_2 | 37.665 | RH_Cont_PFCmp_1 | 12.831 |
| RH_Cont_PFCl_8 | 32.693 | RH_Cont_PFCv_1 | 11.117 |
| RH_Cont_Par_1 | 29.967 | RH_Cont_PFCl_5 | 11.085 |
| RH_Cont_PFCmp_1 | 27.51 | RH_Cont_Par_2 | 10.502 |
| LH_Cont_pCun_2 | 23.14 | RH_Cont_Par_6 | 10.5 |
| RH_Cont_Cing_2 | 21.408 | LH_Cont_Par_5 | 10.077 |
| RH_Cont_PFCl_7 | 19.665 | LH_Cont_Par_4 | 9.913 |
| RH_Cont_PFCl_4 | 19.567 | RH_Cont_pCun_2 | 9.134 |
| RH_Cont_PFCv_1 | 18.369 | RH_Cont_Par_1 | 8.646 |

| | | | |
|---|---|---|---|
| LH_Cont_pCun_1 | 17.731 | RH_Cont_PFCl_12 | 8.053 |
| RH_Cont_PFCmp_2 | 15.116 | RH_Cont_PFCl_9 | 7.36 |
| LH_Cont_Par_2 | 14.115 | LH_Cont_pCun_1 | 7.188 |
| RH_Cont_PFCl_3 | 12.258 | RH_Cont_PFCl_8 | 6.012 |
| RH_Cont_PFCl_10 | 11.716 | LH_Cont_PFCl_5 | 5.107 |
| RH_Cont_PFCl_13 | 11.688 | RH_Cont_PFCl_11 | 4.805 |
| RH_Cont_PFCl_5 | 11.484 | LH_Cont_Par_6 | 4.721 |
| LH_Cont_Par_3 | 10.568 | LH_Cont_PFCv_1 | 4.138 |
| LH_Cont_PFCl_2 | 10.478 | LH_Cont_PFCl_3 | 3.933 |
| LH_Cont_PFCl_6 | 10.096 | LH_Cont_PFCl_6 | 3.783 |
| LH_Cont_Par_1 | 9.992 | RH_Cont_PFCl_6 | 3.527 |
| RH_Cont_pCun_2 | 9.417 | RH_Cont_Par_3 | 3.475 |
| LH_Cont_Cing_2 | 8.878 | LH_DorsAttn_Post_9 | 10.14 |
| RH_Cont_PFCl_2 | 8.45 | RH_DorsAttn_Post_12 | 7.853 |
| LH_Cont_Par_4 | 8.424 | LH_DorsAttn_Post_4 | 7.695 |
| LH_Cont_Cing_1 | 7.663 | LH_DorsAttn_Post_12 | 6.81 |
| RH_Cont_PFCl_1 | 7.543 | RH_DorsAttn_Post_15 | 5.657 |
| LH_Cont_PFCmp_1 | 6 | RH_DorsAttn_Post_16 | 5.329 |
| RH_Cont_Temp_2 | 4.35 | RH_DorsAttn_Post_14 | 5.272 |
| RH_Cont_Cing_1 | 4.283 | LH_DorsAttn_FEF_2 | 5.002 |
| RH_Cont_PFCl_6 | 3.983 | LH_DorsAttn_Post_7 | 4.545 |
| LH_Cont_Temp_1 | 3.514 | RH_DorsAttn_Post_5 | 4.365 |
| RH_Cont_PFCl_11 | 3.428 | RH_DorsAttn_Post_11 | 3.413 |
| RH_DorsAttn_Post_3 | 26.911 | LH_Default_Par_4 | 18.827 |
| RH_DorsAttn_Post_7 | 25.829 | LH_Default_Temp_6 | 14.856 |
| RH_DorsAttn_Post_15 | 22.526 | RH_Default_pCunPCC_4 | 14.543 |
| RH_DorsAttn_Post_4 | 21.592 | RH_Default_PFCdPFCm_4 | 12.88 |
| RH_DorsAttn_PrCv_1 | 19.134 | LH_Default_Par_6 | 12.447 |
| LH_DorsAttn_Post_15 | 16.96 | LH_Default_PFC_13 | 12.06 |
| RH_DorsAttn_FEF_1 | 15.356 | LH_Default_Par_2 | 12.047 |
| RH_DorsAttn_Post_1 | 13.559 | LH_Default_PFC_17 | 11.74 |
| LH_DorsAttn_Post_2 | 13.064 | LH_Default_Temp_10 | 11.386 |
| RH_DorsAttn_Post_17 | 12.209 | LH_Default_Temp_9 | 11.384 |
| LH_DorsAttn_Post_6 | 12.159 | RH_Default_Par_3 | 11.314 |
| LH_DorsAttn_Post_9 | 11.755 | LH_Default_PFC_20 | 11.177 |
| RH_DorsAttn_Post_11 | 10.478 | LH_Default_PFC_9 | 9.259 |
| LH_DorsAttn_Post_7 | 9.994 | LH_Default_Par_1 | 9.043 |
| LH_DorsAttn_PrCv_1 | 7.961 | LH_Default_Temp_3 | 8.637 |
| RH_DorsAttn_Post_14 | 7.076 | RH_Default_Temp_8 | 8.194 |
| LH_DorsAttn_Post_4 | 6.895 | RH_Default_pCunPCC_5 | 8.155 |
| RH_DorsAttn_FEF_2 | 5.449 | RH_Default_Par_4 | 7.807 |
| LH_DorsAttn_FEF_1 | 4.811 | RH_Default_pCunPCC_8 | 7.355 |
| LH_DorsAttn_Post_3 | 4.5 | LH_Default_pCunPCC_6 | 7.239 |
| RH_DorsAttn_Post_2 | 4.253 | LH_Default_pCunPCC_7 | 7.204 |

| | | | |
|---|---|---|---|
| LH_DorsAttn_PrCv_2 | 4.089 | RH_Default_pCunPCC_3 | 7.202 |
| LH_DorsAttn_FEF_4 | 3.619 | LH_Default_PFC_19 | 6.89 |
| RH_Default_PFCdPFCm_6 | 42.322 | RH_Default_Temp_7 | 6.824 |
| RH_Default_pCunPCC_1 | 36.738 | LH_Default_PFC_24 | 6.726 |
| RH_Default_Temp_3 | 36.262 | RH_Default_PFCdPFCm_7 | 6.709 |
| LH_Default_pCunPCC_1 | 30.207 | LH_Default_PFC_7 | 6.7 |
| RH_Default_Par_4 | 28.72 | LH_Default_PFC_21 | 6.691 |
| RH_Default_PFCv_1 | 25.469 | LH_Default_pCunPCC_2 | 6.576 |
| RH_Default_PFCv_3 | 24.762 | LH_Default_PFC_5 | 6.156 |
| LH_Default_PFC_24 | 24.713 | LH_Default_PFC_12 | 5.951 |
| RH_Default_pCunPCC_3 | 24.159 | LH_Default_Par_7 | 5.724 |
| RH_Default_Par_1 | 23.177 | LH_Default_PFC_14 | 5.66 |
| RH_Default_Temp_1 | 22.195 | LH_Default_Temp_7 | 5.557 |
| LH_Default_Par_6 | 22.032 | RH_Default_pCunPCC_6 | 5.538 |
| RH_Default_Temp_7 | 20.781 | RH_Default_Temp_6 | 5.369 |
| LH_Default_Par_4 | 19.783 | LH_Default_Temp_5 | 5.144 |
| RH_Default_PFCdPFCm_13 | 19.551 | LH_Default_PFC_23 | 4.848 |
| LH_Default_Par_3 | 19.528 | RH_Default_PFCdPFCm_6 | 4.835 |
| LH_Default_pCunPCC_2 | 19.37 | RH_Default_Par_5 | 4.755 |
| RH_Default_PFCv_4 | 19.354 | RH_Default_PFCv_3 | 4.57 |
| LH_Default_PFC_7 | 18.86 | LH_Default_pCunPCC_3 | 4.424 |
| LH_Default_Par_2 | 17.889 | LH_Default_PFC_10 | 4.412 |
| RH_Default_Par_2 | 17.718 | LH_Default_Par_3 | 4.261 |
| LH_Default_PFC_12 | 17.07 | RH_Default_Par_2 | 4.209 |
| RH_Default_Par_3 | 16.422 | RH_Default_PFCv_4 | 4.208 |
| RH_Default_Temp_4 | 14.033 | RH_Default_pCunPCC_1 | 4.185 |
| LH_Default_Temp_5 | 13.285 | LH_Default_pCunPCC_5 | 4.125 |
| LH_Default_Temp_6 | 12.269 | LH_Default_PFC_22 | 3.827 |
| LH_Default_PFC_3 | 11.886 | LH_Limbic_OFC_4 | 13.194 |
| RH_Default_pCunPCC_9 | 11.242 | RH_Limbic_OFC_5 | 10.972 |
| RH_Default_Temp_5 | 11.201 | RH_Limbic_TempPole_4 | 8.512 |
| RH_Default_PFCdPFCm_3 | 11.021 | LH_Limbic_OFC_1 | 7.427 |
| LH_Default_pCunPCC_5 | 10.697 | RH_Limbic_OFC_4 | 6.943 |
| RH_Default_pCunPCC_7 | 10.409 | RH_Limbic_OFC_2 | 6.243 |
| RH_Default_PFCv_2 | 9.729 | LH_Limbic_TempPole_6 | 6.129 |
| LH_Default_PFC_9 | 8.697 | LH_Limbic_OFC_3 | 6.007 |
| LH_Default_PFC_20 | 8.664 | RH_Limbic_OFC_3 | 5.62 |
| LH_Default_pCunPCC_9 | 7.275 | RH_Limbic_OFC_1 | 5.503 |
| RH_Default_Par_5 | 7.189 | RH_Limbic_OFC_6 | 5.157 |
| LH_Default_Temp_9 | 6.634 | LH_Limbic_TempPole_3 | 4.946 |
| LH_Default_PFC_5 | 6.347 | LH_Limbic_OFC_2 | 4.768 |
| LH_Default_Temp_4 | 5.261 | RH_Limbic_TempPole_1 | 4.308 |
| LH_Default_Par_1 | 4.664 | RH_Limbic_TempPole_7 | 4.061 |
| LH_Default_Temp_1 | 4.395 | LH_Limbic_TempPole_7 | 3.561 |

| | | | |
|---|---|---|---|
| RH_Default_pCunPCC_2 | 4.072 | LH_SomMot_9 | 10.015 |
| RH_Default_PFCdPFCm_10 | 3.889 | RH_SomMot_12 | 9.206 |
| LH_Default_Par_7 | 3.638 | LH_SomMot_12 | 6.75 |
| RH_Default_Temp_6 | 3.514 | RH_SomMot_11 | 5.202 |
| LH_Default_PFC_17 | 2.728 | RH_SomMot_14 | 5.068 |
| RH_Limbic_TempPole_4 | 35.384 | LH_SomMot_8 | 5.021 |
| RH_Limbic_OFC_4 | 23.469 | RH_SomMot_21 | 4.546 |
| RH_Limbic_OFC_2 | 20.29 | RH_SomMot_10 | 3.073 |
| RH_Limbic_OFC_6 | 19.337 | RH_SomMot_22 | 2.734 |
| LH_Limbic_TempPole_6 | 19.252 | RH_SalVentAttn_TempOccPar_1 | 19.429 |
| LH_Limbic_TempPole_7 | 16.867 | RH_SalVentAttn_TempOccPar_3 | 11.948 |
| RH_Limbic_TempPole_5 | 14.948 | LH_SalVentAttn_ParOper_2 | 10.732 |
| RH_Limbic_TempPole_7 | 13.985 | LH_SalVentAttn_TempOcc_1 | 9.272 |
| RH_Limbic_TempPole_2 | 13.845 | RH_SalVentAttn_Med_1 | 7.569 |
| LH_Limbic_TempPole_3 | 13.494 | RH_SalVentAttn_TempOccPar_6 | 6.94 |
| RH_Limbic_OFC_5 | 13.454 | RH_SalVentAttn_TempOccPar_2 | 4.856 |
| RH_Limbic_TempPole_1 | 13.247 | LH_SalVentAttn_ParOper_4 | 4.184 |
| LH_Limbic_OFC_2 | 13.142 | RH_SalVentAttn_TempOccPar_7 | 3.599 |
| RH_Limbic_OFC_1 | 12.03 | RH_SalVentAttn_FrOperIns_7 | 2.864 |
| RH_Limbic_TempPole_6 | 11.481 | RH_Vis_19 | 8.369 |
| RH_Limbic_TempPole_3 | 8.797 | LH_Vis_7 | 7.468 |
| LH_Limbic_OFC_4 | 8.691 | RH_Vis_9 | 7.282 |
| RH_Limbic_OFC_3 | 8.684 | RH_Vis_18 | 7.082 |
| LH_Limbic_OFC_1 | 7.799 | LH_Vis_2 | 6.22 |
| LH_Limbic_TempPole_5 | 7.617 | RH_Vis_26 | 5.974 |
| LH_Limbic_TempPole_1 | 7.551 | RH_Vis_2 | 3.985 |
| LH_Limbic_TempPole_4 | 5.945 | LH_Vis_12 | 3.789 |
| LH_Limbic_TempPole_2 | 5.801 | LH_Vis_21 | 3.739 |
| LH_Limbic_TempPole_8 | 3.853 | RH_Vis_6 | 3.734 |
| LH_Limbic_OFC_3 | 3.763 | LH_Vis_20 | 3.641 |
| LH_Limbic_OFC_5 | 3.22 | RH_Vis_5 | 3.516 |
| RH_SomMot_22 | 12.937 | RH_Vis_30 | 3.464 |
| RH_SomMot_17 | 11.688 | LH_Vis_4 | 3.292 |
| RH_SomMot_2 | 11.363 | LH_Vis_16 | 3.261 |
| RH_SomMot_8 | 10.948 | | |
| RH_SomMot_23 | 10.537 | | |
| RH_SomMot_32 | 4.185 | | |
| RH_SomMot_20 | 3.547 | | |
| LH_SomMot_1 | 2.812 | | |
| RH_SalVentAttn_TempOccPar_7 | 39.829 | | |
| RH_SalVentAttn_TempOccPar_3 | 38.305 | | |
| RH_SalVentAttn_PFCl_1 | 36.325 | | |
| RH_SalVentAttn_Med_3 | 22.762 | | |
| RH_SalVentAttn_Med_4 | 22.099 | | |

| | |
|---|---|
| RH_SalVentAttn_PrC_1 | 21.914 |
| RH_SalVentAttn_FrOperIns_8 | 21.372 |
| RH_SalVentAttn_TempOccPar_6 | 20.064 |
| LH_SalVentAttn_ParOper_4 | 18.752 |
| LH_SalVentAttn_FrOperIns_9 | 18.678 |
| LH_SalVentAttn_FrOperIns_1 | 17.409 |
| RH_SalVentAttn_FrOperIns_1 | 15.347 |
| RH_SalVentAttn_FrOperIns_5 | 15.311 |
| RH_SalVentAttn_TempOccPar_1 | 15.29 |
| LH_SalVentAttn_Med_1 | 15.213 |
| LH_SalVentAttn_PFCl_1 | 15.039 |
| LH_SalVentAttn_ParOper_2 | 11.968 |
| RH_SalVentAttn_TempOccPar_2 | 11.958 |
| LH_SalVentAttn_FrOperIns_2 | 11.625 |
| RH_SalVentAttn_FrOperIns_2 | 10.125 |
| LH_SalVentAttn_TempOcc_1 | 10.008 |
| LH_SalVentAttn_FrOperIns_3 | 9.029 |
| RH_SalVentAttn_Med_1 | 8.871 |
| LH_SalVentAttn_FrOperIns_7 | 6.773 |
| LH_SalVentAttn_Med_4 | 6.054 |
| LH_SalVentAttn_ParOper_3 | 3.165 |
| RH_SalVentAttn_Med_5 | 3.042 |
| LH_SalVentAttn_ParOper_1 | 2.816 |
| LH_Vis_20 | 18.826 |
| LH_Vis_2 | 16.255 |
| RH_Vis_9 | 15.175 |
| RH_Vis_2 | 14.633 |
| RH_Vis_13 | 9.82 |
| LH_Vis_31 | 9.06 |
| LH_Vis_26 | 7.826 |
| RH_Vis_10 | 6.78 |
| RH_Vis_24 | 5.936 |
| RH_Vis_16 | 5.459 |
| RH_Vis_17 | 5.436 |
| LH_Vis_7 | 4.702 |
| LH_Vis_8 | 4.076 |
| LH_Vis_30 | 3.934 |
| RH_Vis_1 | 3.743 |

Table S6. The 20 strongest positive and 20 strongest negative correlations between hub score and composite hub score in LC1.

| **ROI Name** | **CON: r** | **ROI Name** | **PRO: r** |
|---|---|---|---|
| LH_Default_Temp_4 | 0.421 | LH_SalVentAttn_FrOperIns_1 | 0.349 |
| LH_SalVentAttn_FrOperIns_1 | 0.421 | LH_Vis_7 | 0.324 |

| | | | |
|---|---|---|---|
| LH_Default_PFC_6 | 0.417 | LH_Default_PFC_6 | 0.303 |
| RH_Limbic_OFC_5 | 0.353 | RH_DorsAttn_Post_5 | 0.288 |
| LH_Limbic_OFC_4 | 0.341 | LH_Default_PFC_11 | 0.264 |
| RH_Limbic_OFC_1 | 0.336 | RH_SomMot_20 | 0.256 |
| LH_Limbic_OFC_1 | 0.323 | RH_SomMot_40 | 0.256 |
| LH_Limbic_TempPole_3 | 0.288 | RH_Cont_PFCl_3 | 0.25 |
| LH_Cont_PFCl_1 | 0.267 | RH_Default_pCunPCC_2 | 0.241 |
| LH_Limbic_TempPole_2 | 0.26 | LH_Default_pCunPCC_3 | 0.24 |
| LH_SalVentAttn_FrOperIns_2 | 0.256 | RH_SalVentAttn_FrOperIns_4 | 0.237 |
| LH_SalVentAttn_FrOperIns_9 | 0.256 | LH_SalVentAttn_Med_3 | 0.232 |
| RH_Limbic_TempPole_3 | 0.25 | RH_Vis_25 | 0.232 |
| RH_SalVentAttn_FrOperIns_1 | 0.25 | LH_Default_pCunPCC_6 | 0.224 |
| LH_SomMot_1 | 0.246 | LH_SomMot_23 | 0.212 |
| RH_Limbic_OFC_4 | 0.243 | LH_Cont_Par_1 | 0.211 |
| RH_Default_PFCv_1 | 0.239 | RH_Default_pCunPCC_5 | 0.211 |
| RH_Limbic_OFC_6 | 0.234 | LH_Vis_20 | 0.209 |
| LH_Default_PFC_1 | 0.228 | LH_Cont_PFCl_1 | 0.209 |
| LH_Default_PFC_5 | 0.227 | LH_Cont_PFCl_8 | 0.204 |
| LH_DorsAttn_Post_8 | -0.208 | LH_SomMot_8 | -0.205 |
| RH_SalVentAttn_Med_8 | -0.21 | RH_Cont_PFCl_9 | -0.209 |
| LH_Cont_Par_3 | -0.223 | RH_DorsAttn_FEF_2 | -0.213 |
| LH_SomMot_15 | -0.223 | LH_SomMot_20 | -0.215 |
| LH_SomMot_18 | -0.226 | LH_Vis_15 | -0.216 |
| LH_Default_Temp_2 | -0.228 | LH_SomMot_22 | -0.226 |
| LH_Default_Temp_9 | -0.229 | LH_SomMot_18 | -0.227 |
| RH_Vis_9 | -0.236 | LH_Vis_26 | -0.228 |
| RH_Vis_12 | -0.24 | LH_SomMot_10 | -0.233 |
| RH_DorsAttn_Post_6 | -0.244 | LH_Default_Temp_9 | -0.236 |
| LH_SomMot_21 | -0.244 | LH_Cont_Par_3 | -0.238 |
| LH_SalVentAttn_ParOper_1 | -0.25 | LH_DorsAttn_FEF_3 | -0.239 |
| RH_Default_Temp_8 | -0.268 | RH_SomMot_26 | -0.244 |
| LH_Vis_26 | -0.269 | RH_Default_Temp_8 | -0.249 |
| RH_SalVentAttn_TempOccPar_7 | -0.279 | LH_SalVentAttn_ParOper_1 | -0.251 |
| LH_Vis_15 | -0.279 | RH_SomMot_30 | -0.262 |
| LH_Vis_25 | -0.284 | RH_SalVentAttn_Med_8 | -0.268 |
| LH_DorsAttn_Post_11 | -0.287 | RH_DorsAttn_Post_6 | -0.281 |
| LH_Vis_10 | -0.324 | RH_Default_Par_4 | -0.292 |
| RH_SomMot_19 | -0.351 | RH_SomMot_19 | -0.399 |

Table S7. The 20 strongest positive and 20 strongest negative correlations between hub score and composite hub score in LC2.

| **ROI Name** | **CON: r** | **ROI Name** | **PRO: r** |
|---|---|---|---|
| RH_SomMot_20 | 0.307 | LH_Default_Par_3 | 0.332 |

| | | | |
|---|---|---|---|
| LH_DorsAttn_Post_16 | 0.297 | RH_Default_Par_2 | 0.3 |
| RH_SomMot_27 | 0.293 | RH_Vis_10 | 0.29 |
| LH_SomMot_30 | 0.274 | LH_SomMot_7 | 0.255 |
| LH_Vis_27 | 0.238 | RH_Default_Temp_4 | 0.229 |
| LH_SomMot_35 | 0.233 | RH_DorsAttn_Post_1 | 0.228 |
| RH_SomMot_33 | 0.228 | LH_DorsAttn_FEF_2 | 0.225 |
| RH_DorsAttn_Post_9 | 0.227 | LH_DorsAttn_PrCv_1 | 0.221 |
| LH_DorsAttn_Post_12 | 0.225 | RH_Default_Temp_7 | 0.211 |
| LH_DorsAttn_Post_11 | 0.219 | LH_Default_PFC_23 | 0.207 |
| RH_SomMot_23 | 0.219 | RH_Default_pCunPCC_8 | 0.206 |
| LH_Default_PFC_18 | 0.215 | LH_SalVentAttn_FrOperIns_9 | 0.199 |
| LH_SomMot_21 | 0.207 | RH_Vis_2 | 0.198 |
| LH_SomMot_11 | 0.207 | RH_SalVentAttn_FrOperIns_8 | 0.197 |
| LH_Default_Par_6 | 0.207 | LH_Vis_20 | 0.192 |
| RH_Default_PFCdPFCm_13 | 0.206 | LH_Default_pCunPCC_11 | 0.191 |
| RH_SomMot_11 | 0.205 | LH_DorsAttn_Post_4 | 0.191 |
| LH_DorsAttn_Post_8 | 0.2 | RH_Cont_Temp_2 | 0.185 |
| LH_Default_PFC_17 | 0.2 | LH_SomMot_33 | 0.175 |
| RH_DorsAttn_Post_8 | 0.198 | RH_Vis_3 | 0.175 |
| RH_Vis_16 | -0.176 | LH_SomMot_36 | -0.186 |
| RH_SomMot_1 | -0.178 | RH_SomMot_27 | -0.187 |
| RH_Vis_1 | -0.179 | RH_SalVentAttn_TempOccPar_6 | -0.195 |
| RH_Vis_9 | -0.184 | LH_SomMot_28 | -0.195 |
| LH_Vis_18 | -0.186 | RH_SomMot_23 | -0.197 |
| LH_Limbic_TempPole_6 | -0.19 | LH_SomMot_31 | -0.2 |
| RH_DorsAttn_Post_2 | -0.194 | LH_Default_pCunPCC_8 | -0.201 |
| RH_Default_pCunPCC_8 | -0.194 | RH_DorsAttn_Post_8 | -0.205 |
| RH_Vis_5 | -0.196 | RH_Default_Par_1 | -0.206 |
| LH_DorsAttn_Post_4 | -0.209 | RH_Default_Par_5 | -0.207 |
| RH_Vis_6 | -0.211 | RH_Default_pCunPCC_6 | -0.207 |
| RH_Default_Par_2 | -0.213 | RH_Default_Par_3 | -0.214 |
| RH_Vis_23 | -0.223 | RH_DorsAttn_Post_12 | -0.226 |
| LH_SomMot_7 | -0.232 | LH_SalVentAttn_ParOper_1 | -0.229 |
| LH_Vis_8 | -0.244 | RH_Vis_27 | -0.23 |
| LH_DorsAttn_Post_2 | -0.258 | RH_Vis_15 | -0.252 |
| LH_Vis_26 | -0.286 | RH_Limbic_TempPole_1 | -0.259 |
| LH_Default_Par_3 | -0.288 | LH_SomMot_21 | -0.27 |
| RH_DorsAttn_Post_1 | -0.309 | LH_Vis_27 | -0.29 |
| RH_SalVentAttn_TempOccPar_1 | -0.309 | RH_SomMot_20 | -0.3 |

Supplementary Figures

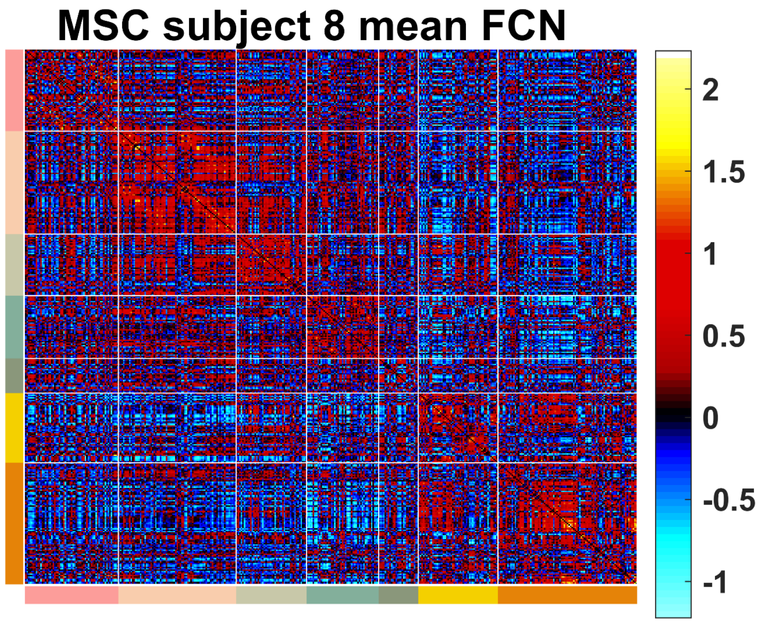


Figure S1. Average functional connectivity of the eighth subject in the MSC database during ten resting state scans.

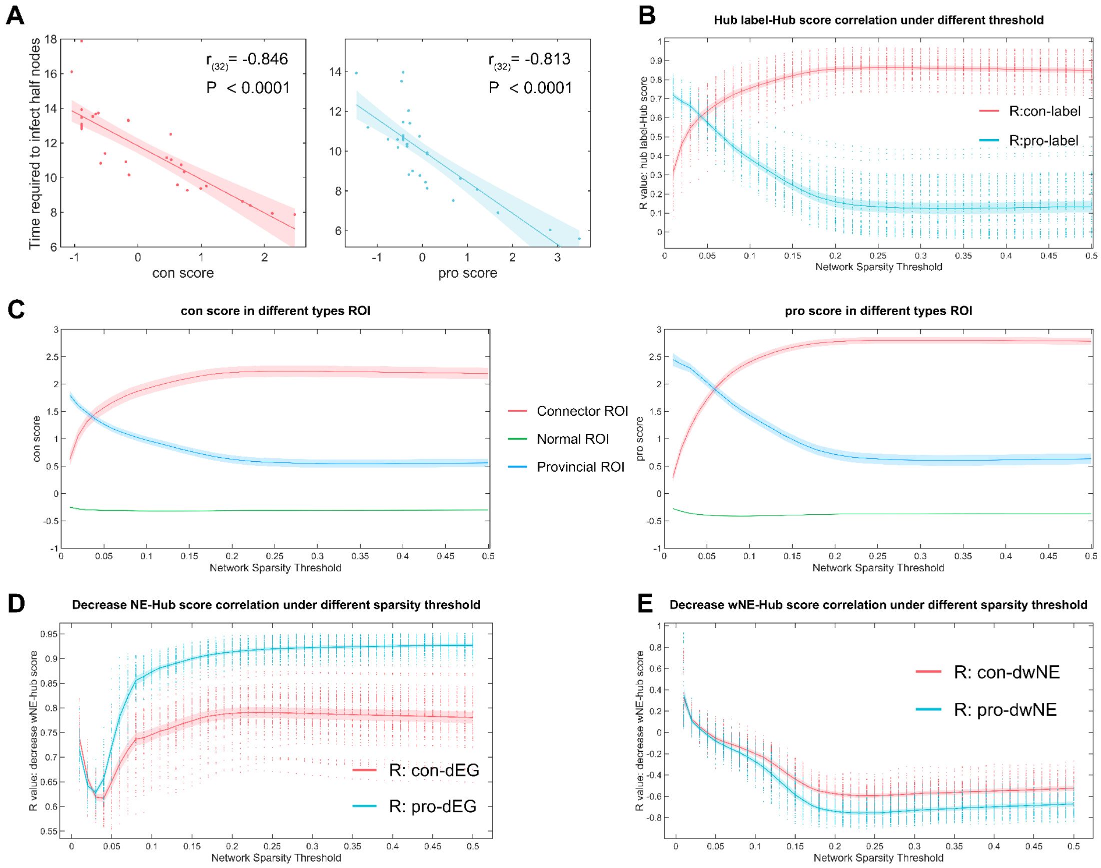


Figure S2. Results of verification using MQGA. (A) Pearson correlation results of the time t taken to diffuse half of the nodes and the hub score. Note: In the propagation simulation of PRO, only propagation to all nodes in the sub-network is considered. (B) Hub score and node label correlation results. (E) Distribution of hub scores of different types of nodes. (C) Distribution of hub scores of different types of nodes.

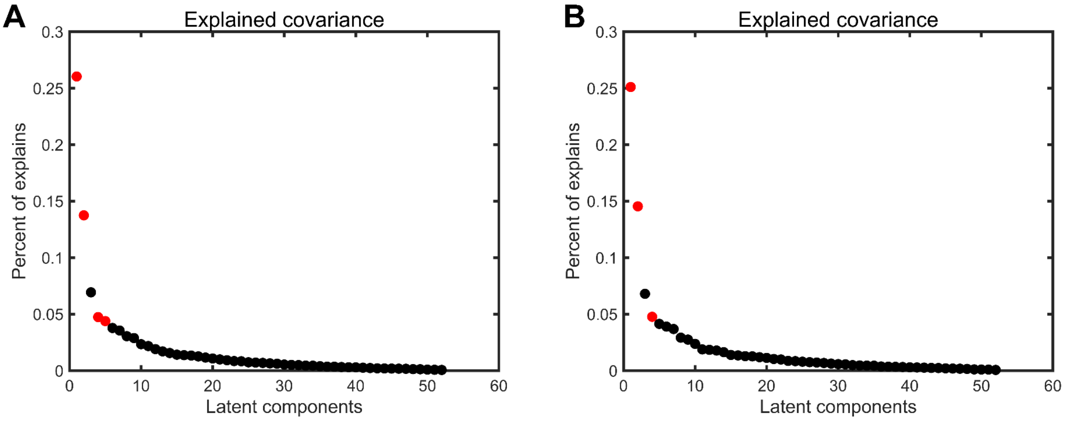


Figure S3. The amount of covariance explained by each LC. (A) PLS analysis based on CON and 52 behavioral indicators. (B) PLS analysis based on PRO and 52 behavioral indicators.

### Reference


1. Gordon, E. M. *et al.* Precision Functional Mapping of Individual Human Brains. *Neuron* **95**, 791-807.e7 (2017).
2. Schaefer, A. *et al.* Local-global parcellation of the human cerebral cortex from intrinsic functional connectivity mri. *Cerebral Cortex* **28**, 3095–3114 (2018).
3. Esteban, O. *et al.* fMRIPrep: a robust preprocessing pipeline for functional MRI. *Nat Methods* **16**, 111–116 (2019).
